# Multi-Omic blood analysis reveals differences in innate inflammatory sensitivity between species

**DOI:** 10.1101/2023.11.30.23299243

**Authors:** David J. Gregory, Feifei Han, Peng Li, Marina Gritsenko, Jennifer Kyle, Frank E. Riley, Deborah Chavez, Vania Yotova, Renata H.M. Sindeaux, Mohamed B. F. Hawash, Fengyun Xu, Li-Yuan Hung, Douglas L. Hayden, Ron G. Tompkins, Robert E. Lanford, Lester Kobzik, Judith Hellman, Jonathan M. Jacobs, Luis B. Barreiro, Wenzhong Xiao, H. Shaw Warren

## Abstract

Vertebrates differ greatly in responses to pro-inflammatory agonists such as bacterial lipopolysaccharide (LPS), complicating use of animal models to study human sepsis or inflammatory disorders. We compared transcriptomes of resting and LPS-exposed blood from six LPS-sensitive species (rabbit, pig, sheep, cow, chimpanzee, human) and four LPS-resilient species (mice, rats, baboon, rhesus), as well as plasma proteomes and lipidomes. Unexpectedly, at baseline, sensitive species already had enhanced expression of LPS-responsive genes relative to resilient species. After LPS stimulation, maximally different genes in resilient species included genes that detoxify LPS, diminish bacterial growth, discriminate sepsis from SIRS, and play roles in autophagy and apoptosis. The findings reveal the molecular landscape of species differences in inflammation, and may inform better selection of species for pre-clinical models.

**One-Sentence Summary:** Fundamental differences are present in blood of humans, mice and other species that differ in sensitivity to inflammation.

## Main Text

Much of the phenotype of many diseases, including bacterial sepsis and viral infections such as SARS-COV2, is caused by excessive inflammation that creates local and systemic pathology beyond that caused by the primary injurious mechanism or microbe. Recent work suggests that the outcome of infection within an individual may depend upon the ability to limit damage from pathogen challenge (*1–5*). In most studies, this has been studied by comparisons within a single species. There are few systematic studies on the ways that different species react to the same pathogen or agonist challenge. We also know little about the mechanisms that different species use to handle the tradeoff between the protection offered by innate inflammation and the associated potential induction of secondary damaging pathology.

For decades bacterial lipopolysaccharide (LPS, endotoxin) has been injected into different species to induce and study inflammatory responses that are intended to mimic human inflammation. Although there are substantial differences in preparations and the conditions studied have not been identical, the bioactive moiety of LPS (lipid A) is generally conserved, so that it is possible to construct a rough ordering of the sensitivity of species to LPS (Fig 1A). These data indicate that species differ over a hundred thousand -fold in sensitivity to LPS. Some species, including rodents and some non-human primates used in many disease models, are quite resilient to LPS. In contrast, humans fall on the end of extreme sensitivity. Most studies in humans have utilized a standardized reference endotoxin prepared by the FDA and administered to volunteers at a dose of 2-4 ng LPS/kg, which causes predictable flu-like symptoms, fever and cytokine release (*6*). Two documented cases of endotoxin injected into humans at higher doses (27ng/kg and 17 ug/kg), caused shock and required intensive care support (*7*, *8*).

**Fig. 1.**
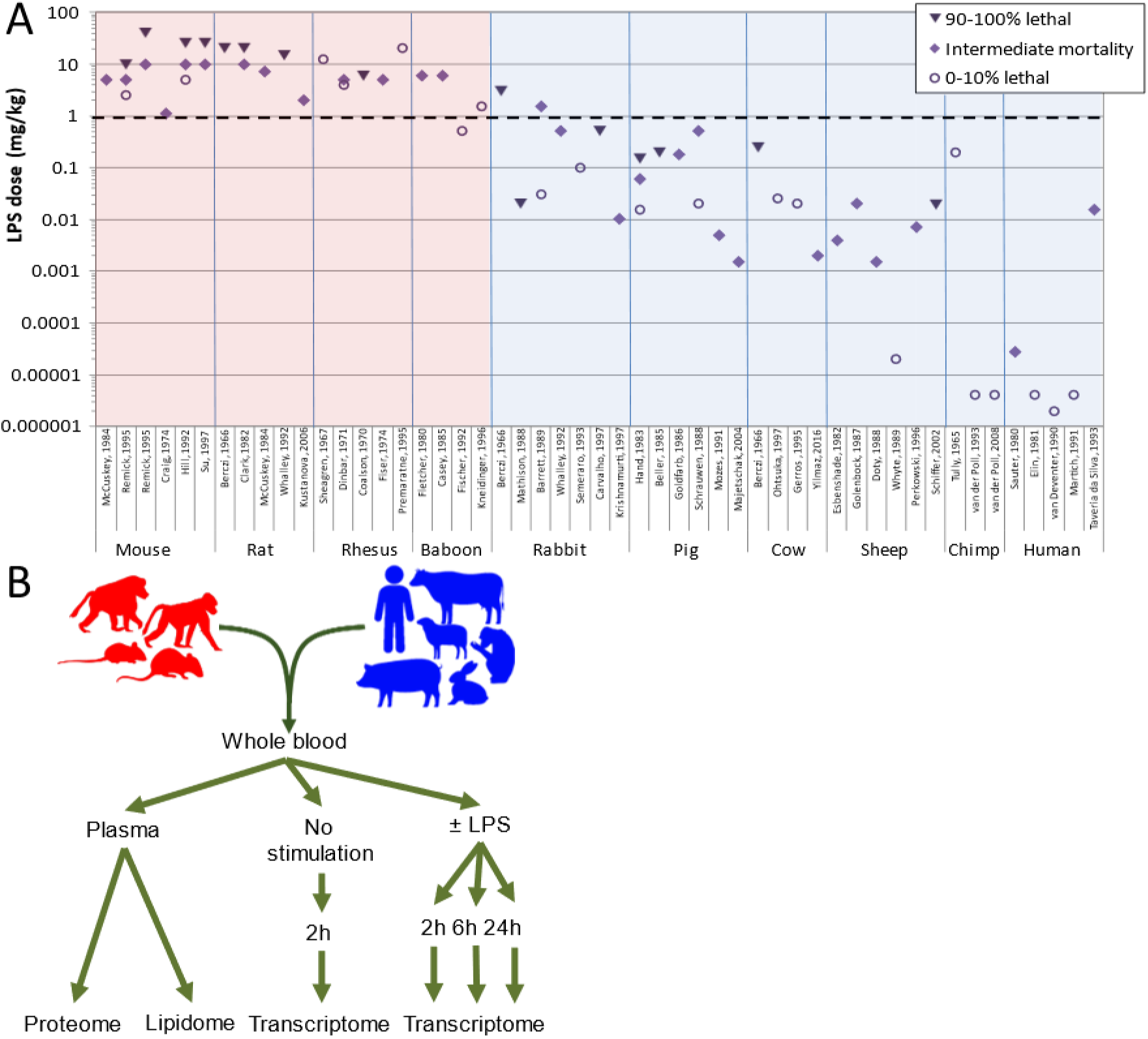
Rationale of study. **(A)** Lethal doses of LPS in different species. Values are selected from representative studies (shown on the x-axis) where LPS was injected i.p. or i.v. and survival reported. Wherever possible, studies using multiple doses of *E. coli* LPS injected as a single bolus are reported. Resilient species, defined here as those that survive a dose of 1 mg/kg body weight (dashed line), are indicated with red background, sensitive species are indicated with a blue background. References are given in Table S1. **(B)** Outline of study. Whole blood from 4-5 individuals of 4 resilient species (mouse, rat, rhesus, and baboon) and 6 sensitive species (rabbit, pig, cow, sheep, chimp, human) was incubated ex vivo with 0, 1, 10, or 100 ng/mL *E. coli* LPS for 2, 6, or 24h followed by leukocyte mRNA sequencing or MS/MS analysis of lipids and proteins was performed on plasma without prior incubation.

To better understand how different species respond to an agonist challenge we compared the proteomics, HDL proteomics, and lipidomics of plasma and the transcriptomics of leukocytes in the whole blood of 10 species that vary widely in sensitivity to in vivo LPS challenge. We used carefully controlled and standardized protocols to collect the plasma and the whole blood of 5 mice, rats, rhesus monkeys, baboons, rabbits, pigs, cows, sheep, chimpanzees, and humans into heparinized tubes containing no or varied doses of LPS. Plasma and leukocytes were harvested at 2, 6, and 24 hours and analyzed for protein or lipid and mRNA (Fig 1B). For the purpose of analysis, we divided species into two groups: those “resilient” to LPS (in vivo sensitivity of > 1mg/kg (mouse, rat, rhesus, baboon) and those “sensitive” to LPS (rabbits, pigs, cows, sheep, chimpanzees, and humans) (Fig 1A). The terminology for underlying resilience to LPS challenge between species is not precise (*1–5*). Baseline resilience to LPS differs from the resilient state induced by prior LPS challenge as well as from the ability to limit the damage caused by a given live parasite burden, both of which have been previously defined as tolerance (*1*, *9*). We use “sensitivity” and “resilience” as the terminology for this article (*10*).

## Results

### Proteomics, HDL Proteomics, and Lipidomics

Plasma proteins condition cellular responses to LPS (*10*). To assess possible protein differences in the plasmas of sensitive and resilient species, we performed an exploratory proteome analysis of pooled plasma samples for each species, as described in Materials and Methods (Fig 2A-C, Fig S1). This approach identified a total of 122 proteins that discriminated between the groups, by differential abundance and/or detection in one group only (Data S1). These included known modulators of inflammatory cell activity, such as components of the complement and coagulation pathways, TGFβ, and lipoproteins. Since high-density lipoprotein is anti-inflammatory and neutralizes LPS (*11*), we purified HDL from each species’ plasma and studied the HDL proteomes using the same analytic approaches. Between 93 and 235 HDL-associated proteins were identified, of which the abundance of 83 discriminated between the sensitive and resilient groups (Fig S2-3 and Data S2). We also identified lipid components in the plasma to determine a species-specific lipidomic profile. There were 520 identified and quantified lipids across the species in 22 subclasses (Fig S4), but there was no clear discernable pattern of lipid differentiation between sensitive and resilient species.

**Fig. 2.**
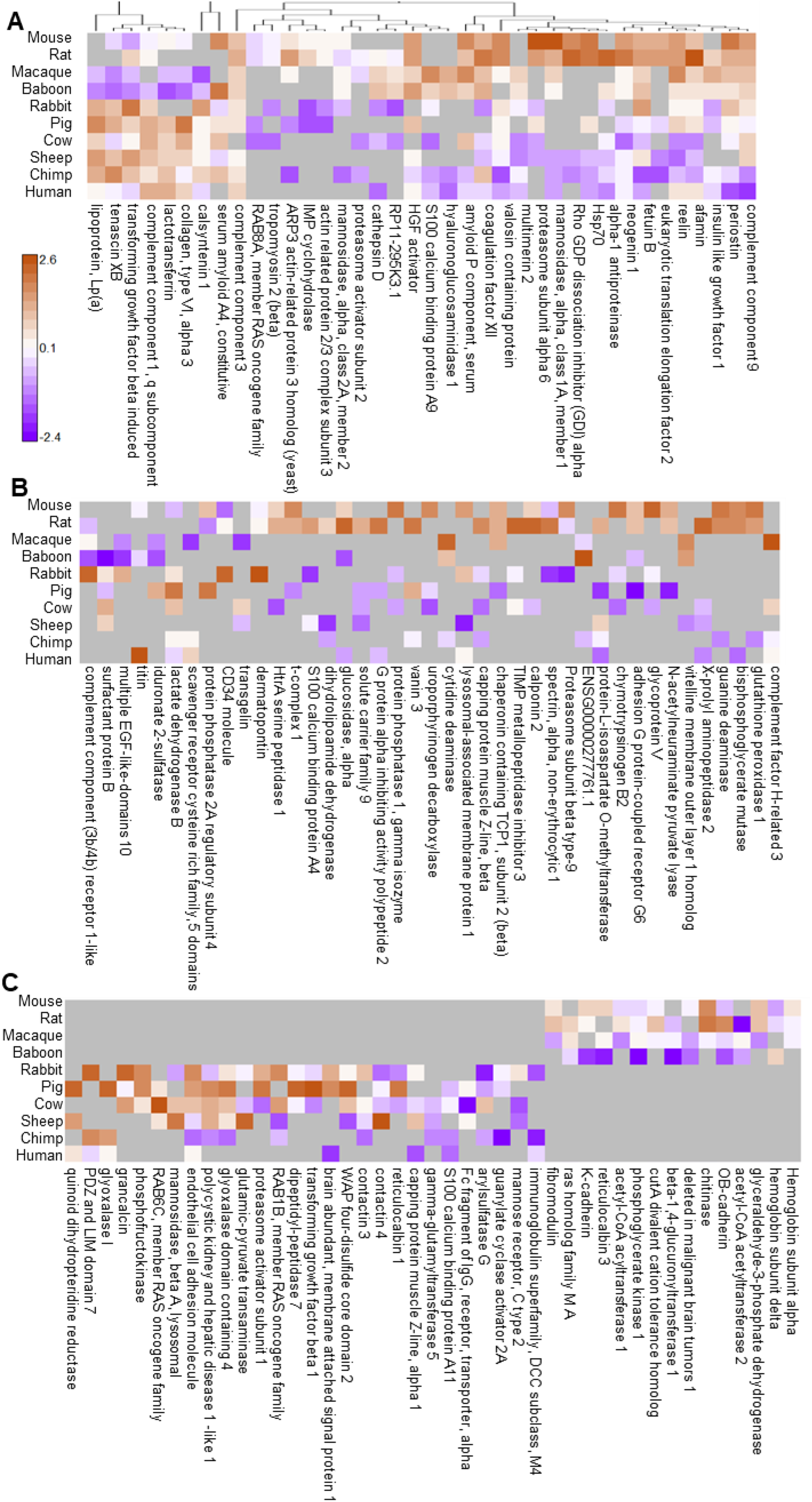
Overview of plasma proteins identified as differentially abundant in comparison of resilient versus sensitive species. Three different comparisons were performed resulting in 122 total proteins identified as differentially abundant. Quantitative protein level values are based upon scaled LFQ intensities combined from peptide level intensities. Color scale represented as scaled quantitative abundance differences with brown representing higher abundance and purple lower abundance for each individual protein. A) Mapping of 38 proteins at *p* <0.05, Pearson correlated. B) Mapping of 41 proteins based upon higher fold-change abundance (+/-3.0 in log2 phase, minimum of 4 occurrences), bimodal correlation. C) Mapping of 43 proteins with yes/no abundance based upon a minimum of 3 species observations, bimodal correlation.

### Transcriptomics at Baseline

We next profiled the gene expression in leukocytes, which are the primary drivers of inflammation, at rest and following LPS stimulation. Our earliest time point provides an estimate of the transcriptomic state of the circulating leukocytes in the blood of the different species, i.e. the baseline state. At baseline, there was a tendency of resilient species to group together and away from sensitive species across the first three principal components (Fig 3A), suggesting that different species and their sensitivity to LPS might be distinguished by their resting gene expression.

**Fig. 3.**
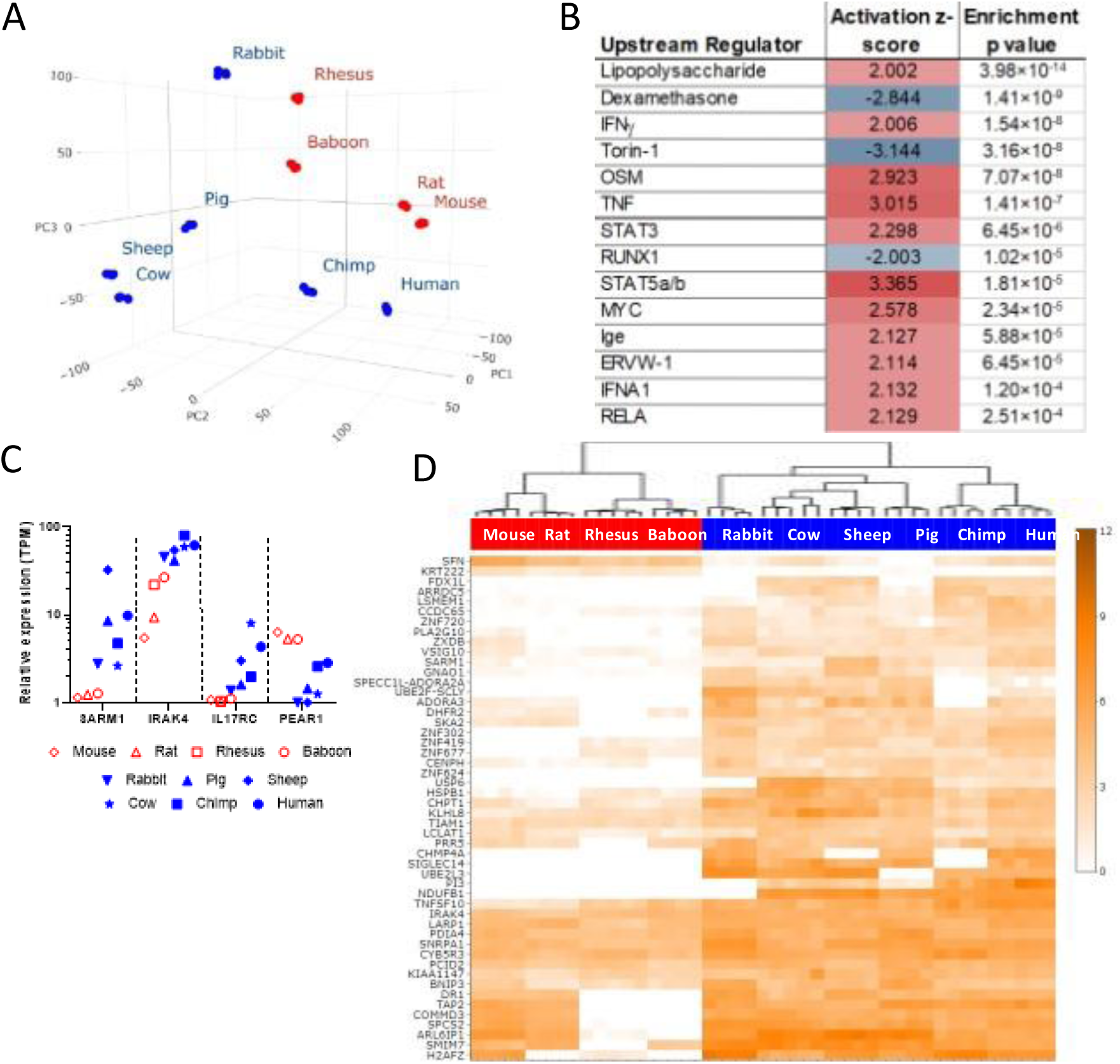
Leukocyte transcriptomes at baseline. **(A)** Principal component analysis showing overall distribution of individual mRNA abundance between individuals. **(B)** Regulators of gene expression that are consistent with the differences between expression profiles of resilient and sensitive species. **(C)** Average expression of four example genes in each species. **(D)** Clustering of the 50 genes with the greatest expression differences between sensitive and resilient species. Color intensity scale: transcript abundance (log_2_ TPM).

We sought to identify transcriptomic features that distinguished sensitive and resilient species. Direct comparison identified 3,012 genes with significantly different expression (2-fold difference, FDR<0.05) between the two groups. Of these, 1,222 were expressed at higher levels and 1790 at lower levels in sensitive species than in resilient species (Table S3). We used Ingenuity Pathway Analysis (Qiagen, Inc) to identify upstream regulators that could mediate this difference (Fig 3B). Strikingly, the most significantly enriched factor with a positive activation z-score was LPS, suggesting that the gene expression profile of resting leukocytes from sensitive animals resembles that of cells that have already been stimulated with LPS when compared to resting leukocytes from resilient animals. The next highest modifiers with a positive activation Z score were IFNγ, Oncostatin M (OSM), TNF and Stat3, which are all potent mediators of inflammation and immunity. The most enriched modifier with a negative z-score, indicating those agents predicted to reduce genes leading to a sensitive gene profile, was dexamethasone, a potent anti-inflammatory agent. The next most enriched regulators are also linked to inflammation: torin-1 is an inducer of autophagy, which antagonizes inflammation (*12*), and RUNX1, which regulates TLR and NF-κB signaling (*13*). These findings suggest that leukocytes in the blood of sensitive animals such as humans, even in the absence of stimulation, have a gene expression pattern consistent with an inflammatory state when compared with leukocytes from resilient animals. The proportion of different leukocyte populations varied by species but did not correlate with the sensitivity of the species to LPS (Fig S5), indicating that the differences in gene expression reflect differences in leukocyte biology or cell state, rather than cell number.

More stringent analysis of the data identified 144 genes with completely divergent expression between the classes, i.e. for which there was no overlap in expression between sensitive and resilient animals with identified orthologs (Data S4). Some examples of possible interest are shown in Fig. 3C, which include IRAK4 which is a known central mediator of the cellular response to LPS and other microbial and inflammatory ligands (*14*), the MyD88 family member SARM1 (*15*), interleukin 17 receptor (IL17RC) and platelet-derived mediator of clotting and inflammation (PEAR1). The 50 protein coding genes with the largest overall differences between the classes and no overlap in expression between sensitive and resistant animals and are shown in Fig 3D. Of note, 49 of these were expressed more strongly in the sensitive species.

### Response to LPS stimulation

We measured cytokine levels in the supernatants of the stimulated whole blood of species for which reagents were available, which included two sensitive species (human and chimpanzee), and four resilient species (baboon, rhesus monkey, mice and rats). Consistent with the in vivo sensitivity profile, the major pro-inflammatory cytokines, IL6 and TNF, as well as G-CSF and IL4, were induced more rapidly and more potently in blood from the sensitive species than the resilient ones (Fig. 4A-D). Similar patterns were found for IFNγ, IL12/23p40, and IL10, but not IL1β (Fig S6). We used targeted proteomics to confirm some of these observations independently of antibody binding (Fig S7).

**Fig. 4.**
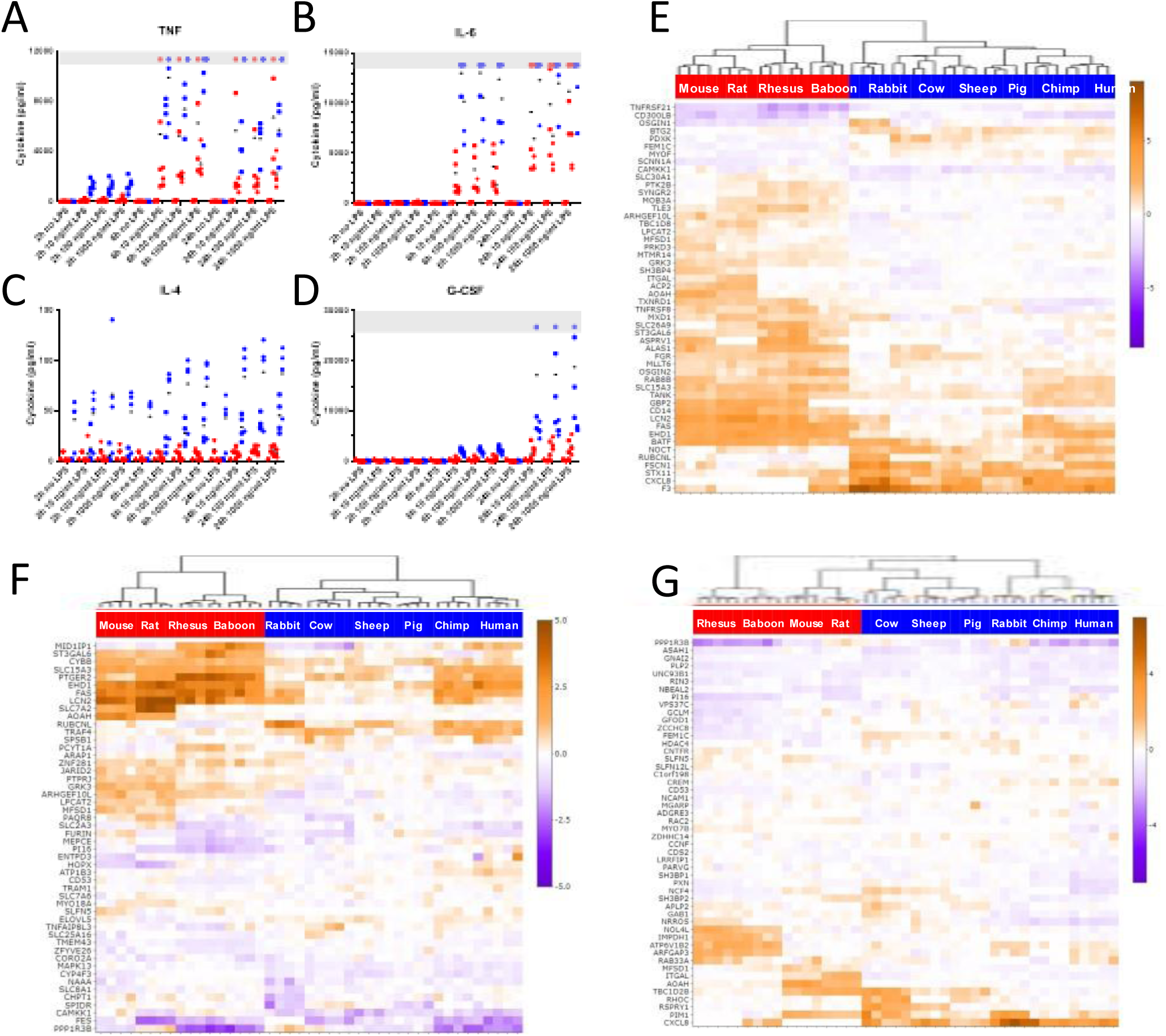
Cytokine and gene expression responses of leukocytes to LPS stimulation. **(A-D)** Cytokine release in a subset of species, measured by Luminex. **(E-G)** Cluster analysis of the 50 genes with the largest difference in response to 10 ng/mL LPS between sensitive and resilient species after 2h, 6h, and 24h, respectively. Color intensity scale: transcript abundance (log_2_ TPM).

The effect of LPS treatment on transcription was studied as the fold change for each transcript relative to unstimulated controls (Fig S8A-C). Within each timepoint, individual animals grouped closely with one another, but there was no clear pattern between sensitive and resilient species. The number of differentially expressed genes varied between the 10 species (2-fold changes, FDR<0.05), and more genes increased than decreased in response to LPS for every species (Fig S8D). The greatest response, in terms of numbers of genes regulated, was seen at 6h for most species. There was a consistent but minimal effect of dose escalation for 10, 100, and 1000 ng/mL LPS, and 10 ng/mL was chosen for further analyses. Notably, the large difference between sensitive and resilient groups in numbers of genes observed at baseline was not reflected in differences between the groups in the number of genes responding to LPS treatment: responses of only 144, 187, and 97 genes were significantly different between resilient and sensitive species at 2, 6, and 24 hours respectively (1.5 fold difference, FDR<0.05) (Data S5-7). Clustering of the top 50 genes for each time point is shown in Fig 4F-H.

We next identified genes where the responses of all individual resilient animals differed from that of all sensitive ones. At 2, 6, and 24 hour time points, 48, 60, and 28 genes satisfied this condition, respectively (Data S8-10). The ten genes with maximal differences in LPS-induced fold change between sensitive and resistant species after stimulation are shown in Fig 5A. Remarkably, at 2 hours, 8 of these 10 genes, and at 6 hours all 10 genes were induced more strongly in the resilient group, and the protein products of many of these genes are known to play a direct or indirect role in LPS related activities. These include CD14, which is cellular co-receptor for LPS signaling (*16*), AOAH which is an enzyme that detoxifies LPS through removal of fatty acids (*17*), and ST3GAL6 which is a glycosyltransferase that regulates the stability of another LPS detoxifying enzyme, alkaline phosphorylase (*18*) (Fig 5 B-D). Five of these were identified at both 2 and 6 hours (FAS, EHD1, ST3GAL6, ARGEFL10, and AOAH) and only one (LCN2) was significantly different in the baseline analysis. The remaining genes were also notable. Low expression of ARGEFL10 in CD14+ monocytes is associated with increased death from sepsis (*19*), and the list included genes related to anti-bacterial activity (LCN2 (*20*), regulation of TLR4 responses (SLC15A3)(*21*), LPS sensitivity and autophagy (RUBCNL and SLC8A1) (*22*, *23*) and apoptosis (FAS, TNFRSF21, TNFRSF8) (*24*).

**Fig. 5.**
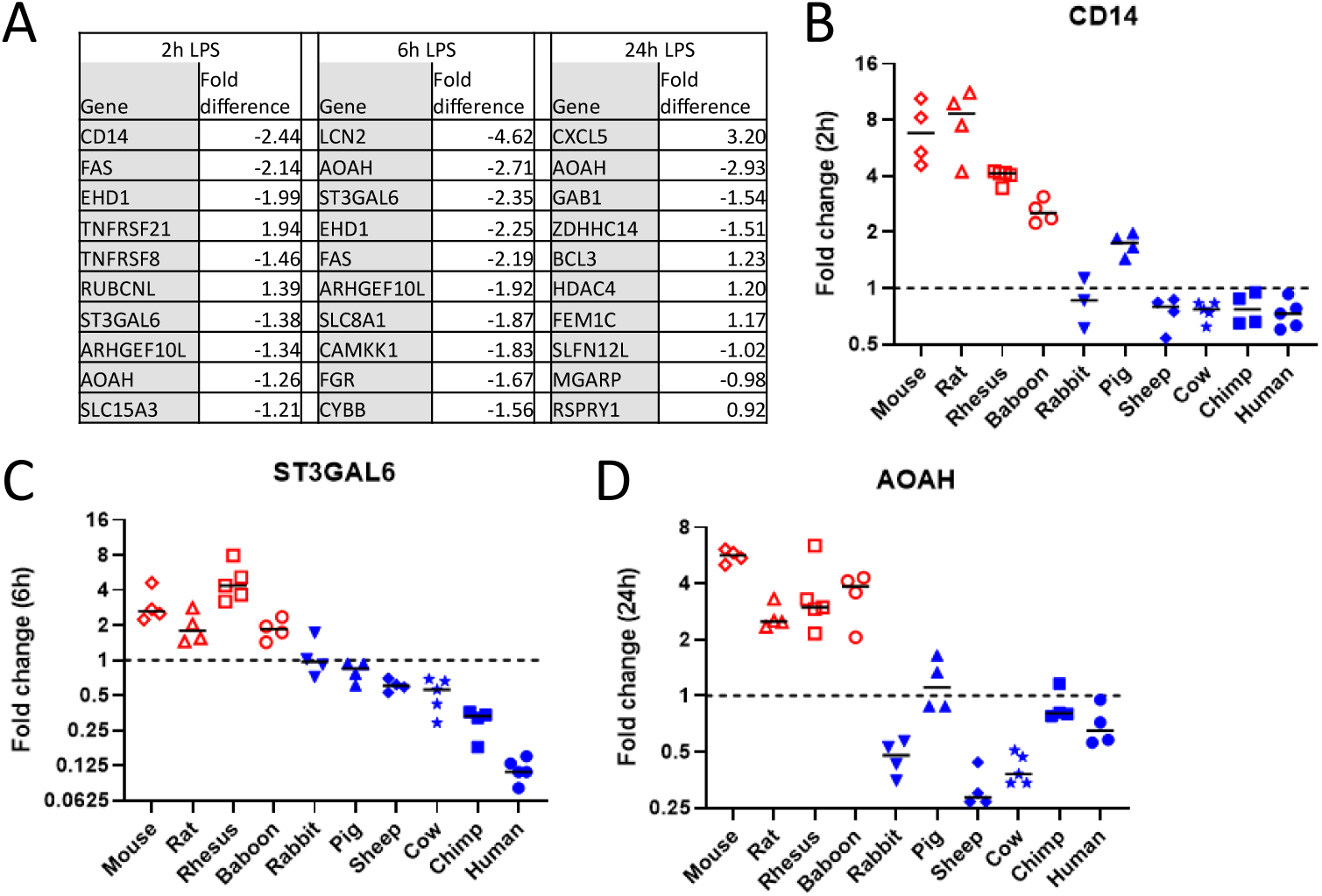
Genes with divergent responses to LPS stimulation. **(A)** The ten genes at each timepoint with the greatest significant (FDR < 0.05) separation in responses between resilient and sensitive animals. Negative numbers indicate genes that are induced less, or repressed more, by LPS in sensitive species than resilient species. **(B-D)** Responses of individual species in example genes from each timepoint. Each point depicts the change in expression (fold change in TPM following incubation with and without 10 ng/mL LPS relative to incubation for the same times in media without LPS) in blood from an individual animal. Bars indicate median fold change for each species. Dashed line indicates no change on stimulation.

## Discussion

LPS is the major driver of inflammation in Gram-negative infections. We hypothesized that there exists a hierarchy of sensitivity to LPS-induced inflammation in different species and that this difference is reflected in blood plasma or leukocyte gene expression. Our findings suggest that information relating to LPS sensitivity of species is contained in circulating blood leukocytes. We found that: 1. species shown in the literature to be sensitive to LPS have an extensive gene activation pattern in whole blood that is suggestive of LPS stimulation at rest (in the absence of LPS stimulation) when compared to the resilient species, and 2. after LPS stimulation, the gene response that most differs between sensitive and resilient species are a relatively small number of genes that are relevant to LPS detoxification, TLR response, bacterial control, and apoptosis. We speculate that the results reflect different paradigms that have evolved to handle microbial exposure, with sensitive species having constitutively active genes at baseline that are prepared to handle infection at the cost of more potentially damaging inflammation, and resilient species activating certain genes after stimulation that can act to limit the damage associated with LPS and inflammation compared to sensitive species.

It is unclear if the classification of the species that we utilized with respect to their sensitivity to LPS is specific to LPS alone or may also apply to other inflammatory stimuli. This question is important given the large numbers of infectious and non-infectious situations in which inflammation is part of a disease process and because LPS is often used as a stimulant to model inflammation beyond Gram-negative infection. There are no other pro-inflammatory agonists reported in the literature that have been administered in sufficient numbers to permit a similar comparison based upon in vivo sensitivity. In humans, the in vivo transcriptomic response to LPS is similar in many ways to that induced by trauma or burns (*25*). The baseline data most likely reflects genes poised to influence the underlying physiological responses to LPS that are sometimes seen minutes after exposure. Stringent analysis of the genes which differ at baseline between sensitive and resistant species revealed numerous genes involved in inflammation, including IRAK4 which was constitutively expressed in the sensitive species relative to the resistant species and which is an important non-specific central mediator of TLR and IL1 stimulation. We did not find obvious baseline genes that were specific to LPS. While the top hit in the pathway analysis was for LPS, most of the other identified pathways (such as interferon gamma) were non-specific.

In contrast to the baseline results, stringent analysis of genes that were maximally increased or decreased with no overlapping between the two groups two or six hours after LPS stimulation resulted in a list of genes which were mostly increased in the resilient species and which are known to be important in either LPS detoxification or clearance, inflammation, and apoptosis. There was a striking difference in directionality between the genes that had maximum differences between two groups at baseline and after stimulation. Baseline genes that most differed between the two groups were almost exclusively increased in the sensitive group, whereas genes that most differed after LPS stimulation were mostly increased in the resilient group. The factors leading to the physiological responses at different times after LPS stimulation are complex, and it is unclear if these genes or their gene products directly mediate or compensate for pathological inflammation.

Immune cells are conditioned by exposure to the surrounding plasma (*10*), and LPS is rapidly detoxified by binding to HDL (*11*), raising the possibility that non-cellular blood components might be related to the species differences in LPS. However, while analysis of the resting plasma proteome, HDL proteome, and lipidome revealed that there were substantial differences between the ten species, we were unable to detect differences between the sensitive and resilient groups that would provide a clear and unifying explanation for the difference in LPS sensitivity in the different species.

There are limitations to our study. The ordering of the sensitivity to the species is based on studies using LPS from different strains of bacteria and different outcome measurements which could have introduced artifacts in the species grouping of responses to LPS. Transcriptomes of all species were related to the human transcriptome. While this was necessary for allowing cross-species comparisons, it can distort the apparent expression of particular genes where there is not 1:1 conservation between species pairs, for example where no ortholog has been identified one or other species, or where a single human gene is represented by multiple orthologs in another species, or vice versa. The species have differing numbers and proportions of types of leukocytes in their whole blood, so that the studies reflect the net sum of all gene responses in the blood. Although the percentages did not correlate with the species grouping, specific conclusions could be confounded by different cells playing a role in the underlying mechanisms. In addition, our results reflect the blood compartment only, which does not necessarily reflect results from different tissues. For technical reasons and to ensure that all results between species were strictly comparable, our earliest time point was at two hours. It is possible that stresses to the cells during sample collection, incubation, and erythrocyte lysis could have altered the response even in the absence of LPS. The lack of detection of proinflammatory cytokines such as TNF in samples in which there was no LPS added is reassuring in this regard. Our analyses of pathways were performed using a program that is based upon prior published gene associations to analyze gene pathways. By design the results of these and similar programs include literature biases that populate the medical and research literature. Regardless of these limitations, our data support the hypothesis that species can be characterized by those that respond with high sensitivity and those that are relatively resilient (Fig 1A) and indicate that information relating to this division is present in blood leukocytes.

To our knowledge, the work represents the first approach to systematically compare the proteomics, lipidomics and transcriptomics between species that are sensitive or resilient to LPS. Humans are at the most sensitive end of the range of species to LPS challenge. The evolutionary advantage for this is not clear, especially considering that many non-human primates are highly resilient, and that their innate immune baseline state does not seem to be associated with any obvious disadvantage. There are however several important implications. First, if innate inflammation is essential for immune defense but at a potential cost of increased inflammatory secondary tissue damage, one might expect humans to be more resilient to infections but also more likely to have inflammatory consequences such as worse symptoms or secondary damage during infection than more resilient species such as mice or rhesus macaques or baboons. Second, our results suggest that the choice of a species to mimic any individual inflammatory disease in humans might be improved by tailoring the species to one that mimics humans in relevant disease specific genes. Our data could be used to choose a species in which to model drugs that have a specific gene target (by matching responses with the human response for that gene). Third, it may be possible to utilize these data as a means of discovering new drug targets or as the basis of a new approach to therapeutic modulation of immunity: Inducing a state similar to those in resilient species in order to temporarily suppress damaging inflammation during infection or other damaging pro-inflammatory conditions without incurring broader immunosuppression.

## Data Availability

All data produced in the present study are available upon reasonable request to the authors. Sequencing data has been submitted to GEO and will be released publicly on formal publication.

## Acknowledgments

We thank Dr. Matthew Hepburn for critical insights throughout this project.

## Author contributions

Conceptualization: DJG, DLH, RGT, REL, LK, JH, JMJ, LBB, WX, HSW

Methodology: FH, PL, MG, JK, FER, DC, VY, RS, BBFH, FX, LYH, JMJ, JBB, WX

Visualization: DJG, FH, FER, JMJ, WX

Funding acquisition: RGT, REL, LK, JH, JMJ, JBB, WX, HSW

Project administration: FER, DJG, WX, HSW

Supervision: REL, JH, JMJ, LBB, HSW Writing – original draft: DJG, HSW

Writing – review & editing: DJG, RGT, REL, LK, JH, JMJ, LBB, WX, HSW

## Materials and Methods

Non-human primates: Blood samples from chimpanzees, baboons and rhesus macaques were obtained from the colony at the Southwest National Primate Research Center (SNPRC) at Texas Biomedical Research Institute. The animals were cared for in accordance with the Guide for the Care and Use of Laboratory Animals. SNPRC is accredited by AAALAC International. All protocols were approved by the institutional IACUC. Animals were selected to be of approximately the same age and were naïve with regard to experimental procedures. Rhesus macaques were sourced from the SNPRC specific pathogen free colony and were negative for Herpes B Virus, Simian immunodeficiency virus, Simian Betaretrovirus (SRV), Simian T-Cell Lymphotrophic Virus. Animals were group housed in outdoor housing except during recovery from procedures, and animals were sedated prior to obtaining blood samples. Chimpanzee samples were obtained prior to the ban on chimpanzee research. Male animals were used because of the high demand of females as breeders, and because the total number of animals involved in the study was too small to demonstrate a significant difference between sexes.

Humans: Blood was obtained from five male human donors in heparin tubes. The protocol was approved by the University of Texas Health Science Center IRB (IRB# HSC20170139H).

Sheep, Pigs, Cows: During routine veterinary check-ups, 10 mL of blood were collected in heparin tubes from each of five adult female cows, sheep, and castrated pigs.

Mice: Because of the small volume of blood in a single mouse, each individual sample was a composite of blood pooled from 25× 10 week old, male C57/BL6J male mice (Jackson Laboratories). Each sample was prepared on a different day. Blood was drawn by cardiac puncture under deep ketamine-xylazine in accordance with Massachusetts General Hospital IACUC protocol (2003N000329).

Rabbits: A total of five 22-week old male New Zealand White rabbits (Western Oregon Rabbit Co, Philomath, OR) were deeply anesthetized with inhaled isoflurane. Then, using sterile technique, the maximum attainable volume of blood was collected by cardiac puncture in accordance with University of California San Francisco IACUC protocol AN152939. Each individual sample was the blood from one rabbit.

Rats: A total of twenty 14-week old male Sprague-Dawley rats (Simonsen Laboratories, Gilroy, CA) were deeply anesthetized with inhaled isoflurane. Using sterile technique they underwent a small thoracotomy incision to expose the heart. Then, using sterile technique, the maximum attainable volume of blood was collected by cardiac puncture in accordance with University of California San Francisco IACUC protocol AN152955. Each individual sample was a composite of blood pooled from 4 rats.

### *Ex vivo* stimulation

In all cases, blood was collected into heparin tubes using meticulous pyrogen-free conditions. TruCulture tubes (Myriad RBM) containing either cell culture media only (“control”) or cell culture media plus 10 ng, 100 ng, or 1 μg/mL *E. coli* 0113 US national reference strain LPS (*26*) (gift of Anthony Rudbach) were prepared in advance, stored at −20 °C or below, and thawed at room temperature immediately prior to use. 1 mL blood was added to each tube and samples were incubated for 2, 6 and 24 h at 37 °C with 5% CO_2_. After incubation, blood in TruCulture tubes was separated according to the manufacturer’s instructions with additional centrifugation at 800 ×g for 10 minutes. Supernatants were saved and frozen at −80 °C for cytokine analysis. The cell pellets were lysed using red blood cell lysis solution for 10 min followed by centrifugation at 2,000 ×g for 3 minutes, and remaining white blood cells were lysed in Qiazol and frozen at −80 °C until library construction (Qiagen). An additional aliquot of blood was immediately separated by centrifugation, without incubation in TruCulture tubes, and plasma stored at −80 °C for proteomic analysis.

### Cytokine analysis

Plasma from mice and rats was assessed for the cytokines reported by Luminex technology (MAGPIX, Millipore Sigma), using commercially available beads (mice MCYTOMAG-70K-13, rats RECYTMAG-65K-12, non-human primates PRCYTOMAG-40K-13, humans HCYTOMAG-60K-13, Millipore-Sigma).

### RNA sequencing

RNA isolated from white blood cells (Qiazol, Qiagen) was quality controlled using A260/A280 ratio, 28S/18S rRNA ratio, and an RNA integrity summary score (Agilent RIN). Total RNA samples (250ngs, RIN>=7.0) were enriched for mRNA, fragmented, and converted into indexed cDNA libraries according to Illumina TruSeq Stranded mRNA protocol and quantity controlled using Agilent TapeStation. Samples were sequenced to at least 25M of 2 x 50bp paired-end reads and assessed using in-house QA/QC metric (Q2Solutions Expression Analysis).

### Gene expression analysis

Sequencing quality was assessed additionally using FASTQC (v0.11.9). Transcriptome annotations of each species were downloaded from Ensembl (https://www.ensembl.org, v90). For each sample, firstly, reads were aligned to human genome hg38 using STAR (*27*) in the established bulk-RNA sequencing workflow of Bcbio (v1.2.1). Secondly, the abundance of the annotated genes of the species was estimated from the RNA-seq reads as Transcripts per Million (TPM) using Sailfish (*27*, *28*). Thirdly, to compare the gene expression between species, the abundance of the genes in each species was then translated to that of the corresponding human orthologues. Here, only genes in a non-human species that had known orthologous human genes (Ensembl v90) were considered in further analysis. When m genes in a non-human species had n orthologous genes in human (m>=0, n>=1), the abundance of each of the human orthologues is assumed to be the sum of the abundance of the m genes divided by n.

### Statistical analysis of the transcriptome data

The Limma package in R (*29*) was utilized to identify transcriptome features significantly different between sensitive and resilient species. RNA sequencing data were analyzed after quantile normalization. For the comparison of baseline expression levels between the two groups of sensitive and resilient species, a mixed effects model was used (*30*), and significant genes were identified with a cutoff of fold difference >=2 and a false discovery rate (FDR) <= 0.05. To evaluate the expression changes in response to LPS stimulation in each species, the expression data for each dose of LPS stimulation (10, 100, and 1000 ng/mL LPS) were compared with those of the control condition (no LPS) separately at each of the time points (2, 6, and 24 hours) using a paired analysis, and a cutoff of fold change >=2 and an FDR <= 0.05 was used. For the comparison of the differences in gene response to LPS stimulation between the groups of sensitive and resilient species <u>at each time point,</u> the differences in fold changes between the stimulation (10nl/ml LPS) and the control were first calculated, followed by analysis using a likewise mixed effects model; significant genes between the two groups were identified with a cutoff of the difference in the fold changes >=1.5 and an FDR <= 0.05. In comparisons of all resilient species vs all sensitive species, a single “missing” annotation was tolerated in the more strongly expressed or induced class, to accommodate incompleteness in genome annotations. Ingenuity Pathway Analysis (*31*) was used to identify changes in activity of upstream regulators, which may help explain the observed difference in gene expression between sensitive and resilient species. The uncorrected, Fisher’s Exact Test P-value for the overlap and Activation Z-score were calculated as previously described (*32*).

### Global plasma proteome analysis

Plasma samples for proteome analysis were collected simultaneously with leukocytes for transcriptome analysis. 30 µL plasma from each animal was pooled (150 µL total) and processed through reduction and tryptic digestion to the peptide form as previously described (*33*). To avoid introducing bias, no affinity depletion was utilized in this study. Instrument analysis was performed using a platform based upon liquid chromatography mass spectrometry (LC-MS) to globally identify and quantify plasma peptide and their corresponding proteins within each sample and species as previously described (*34*). Data generated was based upon label free peak intensity utilizing the MaxQuant data analysis pipeline as previously described (*35*) providing protein level quantitative data (LFQ values) for subsequent comparisons. Data from each unique species LC-MS analysis was searched independently from their respective RefSeq species and subsequently mapped onto a universal human protein for direct species comparison as previously denoted.

### Statistical analysis of the proteome data

Determination of discriminating proteins between sensitive and resilient species was based upon statistical comparisons utilizing the generated protein level quantitative values (LFQ) and utilized the DAnTE InfernoRDN analysis tool (*36*). Specifically, each comparison was comprised of three mutually exclusive approaches. Protein abundances were first subjected to ANOVA analysis for generation of a p value, from which a <0.05 threshold was utilized for an initial capture of discriminating proteins of interest. Secondly, proteins which did not have sufficient replicate abundances to generate a p value were compared via a fold-change abundance threshold (+/-3.0 in log2 phase, minimum of 4 occurrences) resulting in capture of discriminating proteins of interest. Finally, proteins for which we unable to generate a fold-change value, i.e. those which were present/absent in either the sensitive or resilient group, were captured and termed discriminating if they were detected in a minimum of 3 species.

### HDL proteome analysis

Isolation of HDL proteins from plasma for LC-MS analysis followed previously published protocols (*37*). Initial starting amount was ∼300 µl of plasma in biological triplicate. Following density gradient and ultracentrifugation steps, and removal of the HDL containing layer, desalting using Dialysis/Amicon 3K ultracentrifugal filters was used to remove KBr, and Bradford Assay was used to determine protein concentration. Remaining protein processing to peptide form and LC-MS analysis and data analysis was performed identical to global as described above.

### Plasma lipidome analysis

For plasma, 50 μL was used for lipid extraction using a modified Folch extraction (*38*), the MPLEx protocol (*39*). Details of the sample processing were as previously described (*40*) to obtain a total lipid extract (TLE) for analysis. TLEs were analyzed as outlined in Kyle *et al.* (*41*) and as previously described (*40*) utilizing a Waters Acquity UPLC H class system interfaced with a Velos-ETD Orbitrap mass spectrometer for LC-ESI-MS/MS analyses. LC-MS/MS lipidomics data were analyzed using LIQUID (Lipid Informed Quantitation and Identification) (*41*) and as previously described (*40*). To facilitate the quantification of lipids, lipids identified from the MS/MS data from each analysis were then aligned based on their identification, *m/z,* and retention time using MZmine 2 (*42*). Aligned features were manually verified and peak apex intensity values were exported for subsequent statistical analysis.

### Targeted proteome analysis

Selective Reaction Monitoring (SRM) was performed on the panel of primate plasma samples as described above. Crude heavy peptides labeled with 13C/15N on C-terminal lysine and arginine were purchased from New England peptides (Gardner, MA). Trypsin digested samples that had been stored at −80°C until use were processed as previously described (*43*). For each sample the digested peptides were diluted to 0.2 µg/µL containing standards at a final concentration of 250 fmol/µL for 11 protein standards and 500 fmol/µL for 12 proteins. All the samples were analyzed with a nanoACQUITY UPLC® system (Waters Cooperation, Milford, MA) coupled online to a TSQ Vantage triple quadrupole mass spectrometer (Thermo Scientific, San Jose, CA). The LC-SRM platform was configured and utilized as previously described (*44*). Peptides used were DDKPTLQLESVDPK (IL-1β), VNLLSAIK (TNF), ESLLEDFK (IL-10), FLELAYR

SRM data acquired on the TSQ Vantage were analyzed using Skyline software (*45*). Peak detection and integration were determined based on retention time and the relative SRM peak intensity ratios across multiple transitions between light peptide and heavy peptide standards (*46*). All the data were manually inspected to ensure correct peak assignment and peak boundaries. The peak area ratios of endogenous light peptides and their heavy isotope-labeled internal standards (i.e., L/H peak area ratios) were then automatically calculated by Skyline, and the average peak area ratios from all the transitions were used for quantitative analysis of the samples. For targets that had more than one surrogate peptide, correlation graphs were plotted to verify a strong correlation and ultimately the peptide that had the most sensitive response was selected for obtaining quantitative values.

---

**Fig. S1.**
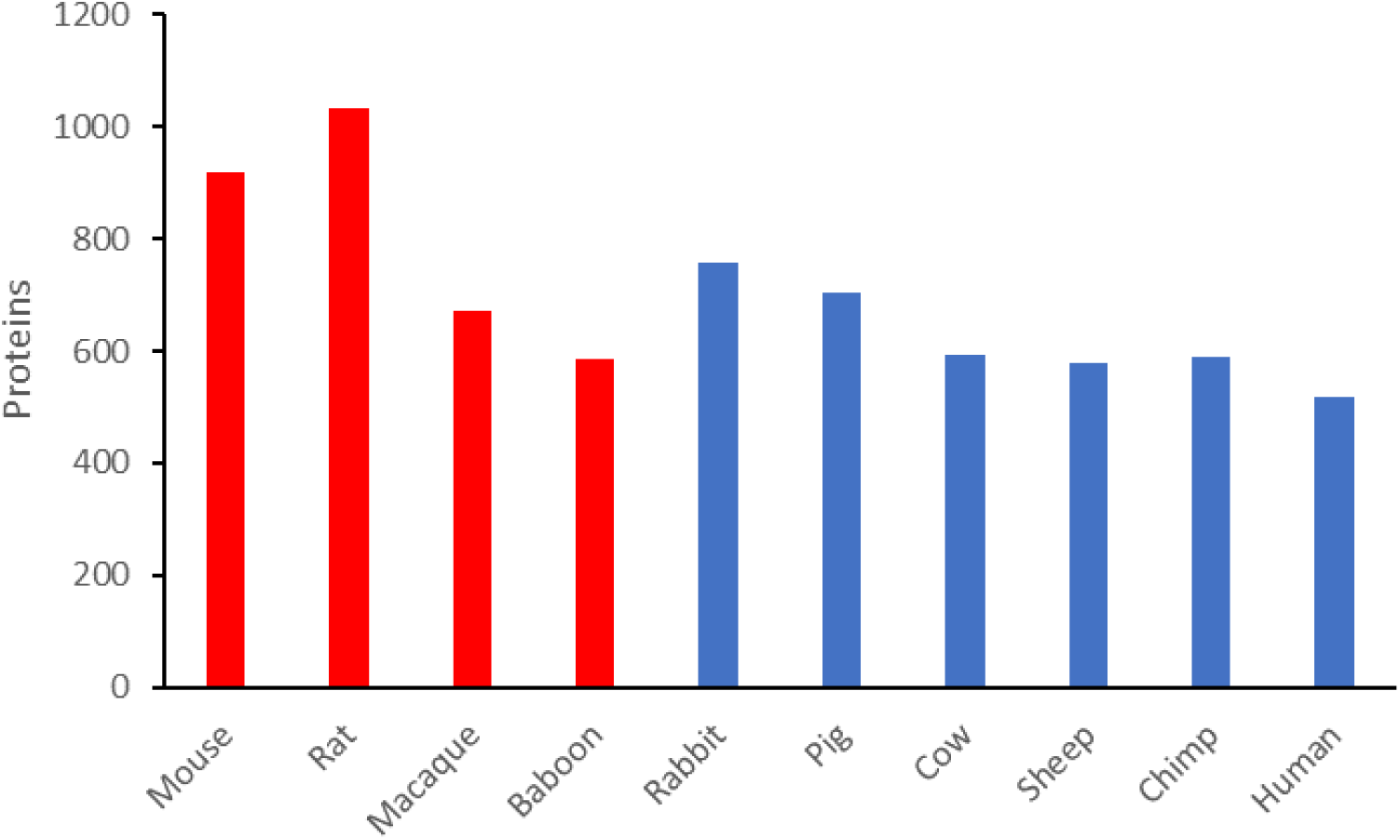
Total number of proteins identified and quantified in whole plasma for each species after LC-MS/MS analysis.

**Fig. S2.**
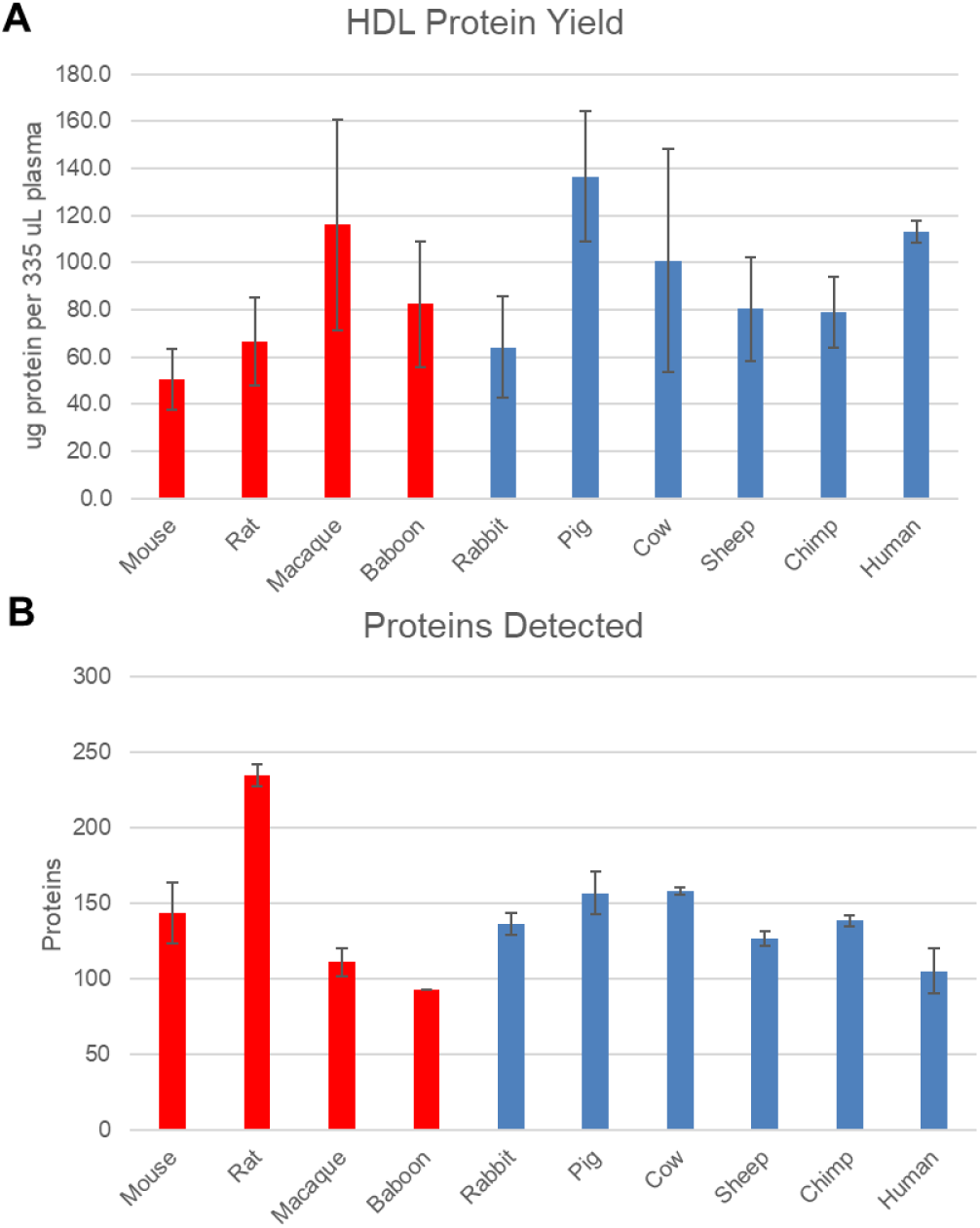
Measured protein yields from HDL protein isolation and detection. A) yield of total protein amount per species after isolation of HDL from replicate plasma samples. B) unique HDL proteins detected and identified per species after LC-MS/MS analysis. Error bars represent biological triplicate analysis.

**Fig. S3.**
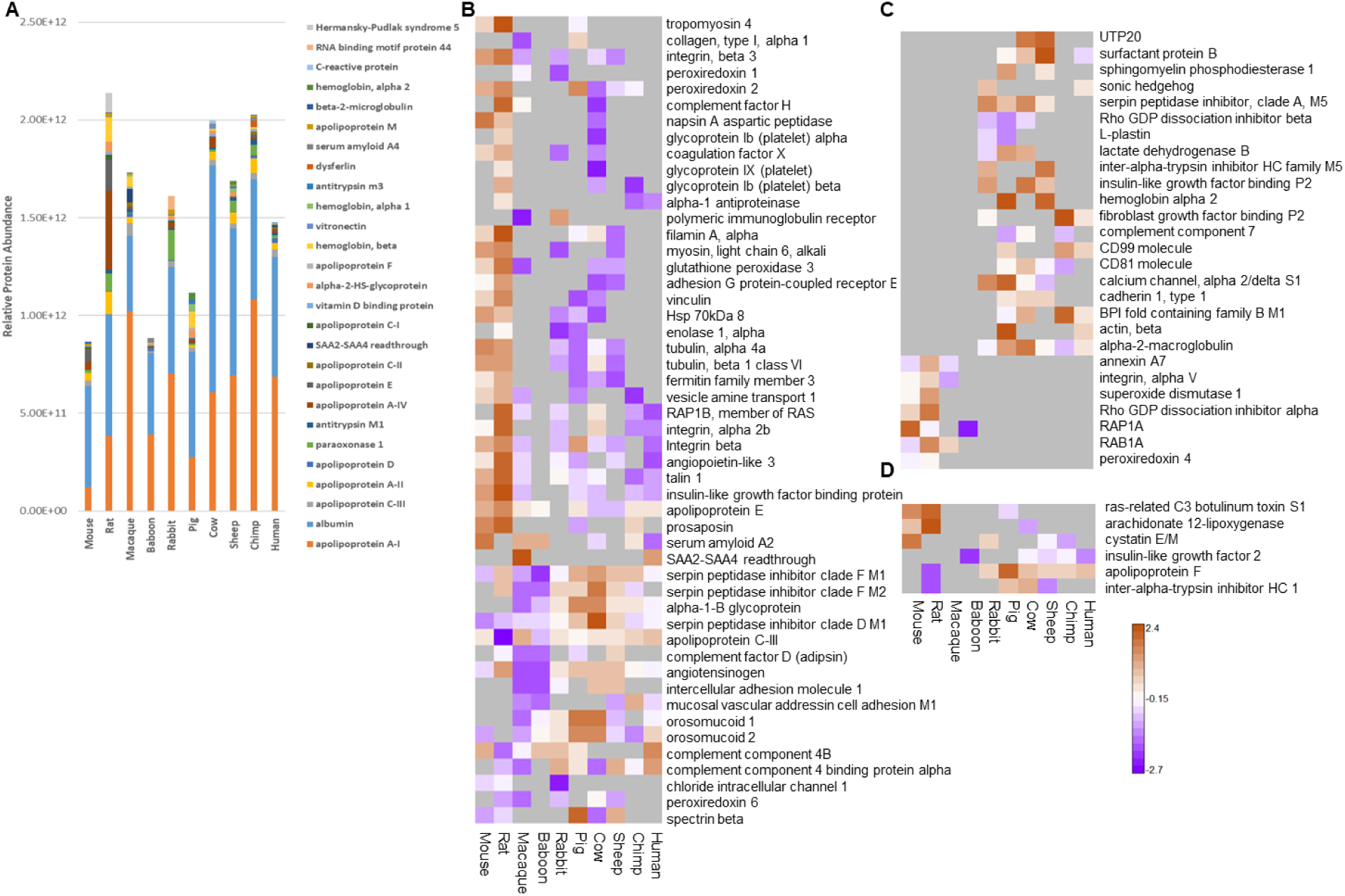
Overview of HDL isolated proteins from plasma and identified as differentially abundant in comparison of resilient versus sensitive species. A) top 10 most abundant HDL proteins per species. Three different comparisons were performed (B-D) resulting in 83 total HDL proteins identified as differentially abundant. Quantitative protein level values are based upon scaled LFQ intensities combined from peptide level intensities. Color scale represented as scaled quantitative abundance differences as shown with red representing higher abundance and purple lower abundance for each individual protein. B) mapping of 50 proteins at *p* <0.05, Pearson correlated. C) mapping of 27 proteins with yes/no abundance based upon a minimum of 3 species observations, bimodal correlation. D) mapping of 6 proteins based upon higher fold-change abundance (+/-3.0 in log_2_ phase, minimum of 4 occurrences), bimodal correlation.

**Fig. S4.**
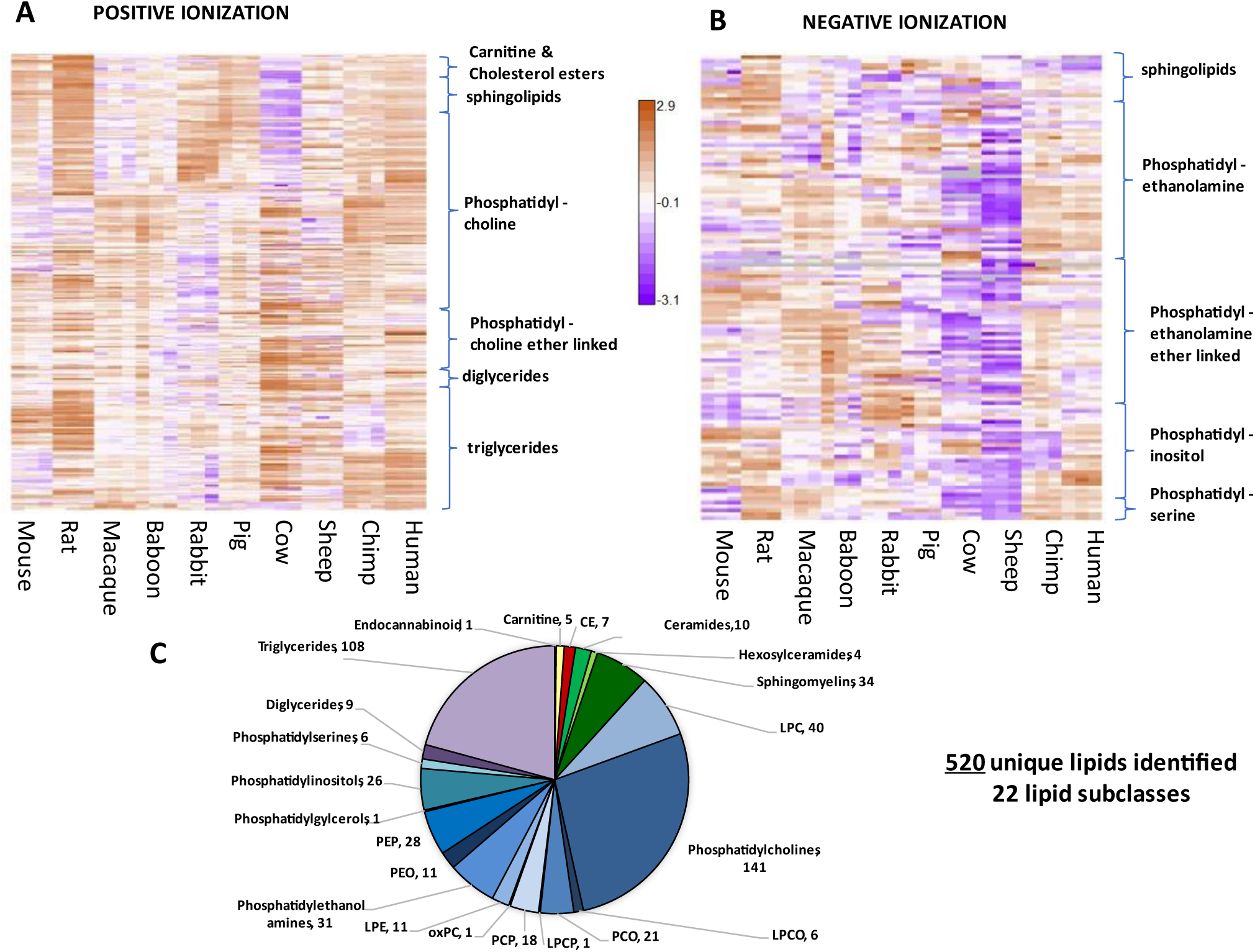
Plasma lipidomes. Abundance of individual lipids that form positive (A) or negative (B) ions detected and quantified via LC-ESI-MS/MS is shown for each of 3 individuals per species, relative to the global mean signal intensity for each lipid. (C) Total number of distinct lipids identified, by lipid subclass. CE: cholesterol esters; LPC: lyso-phosphatidylcholines; LPCO: monoalkylglycerophosphocholines; PCO 1-alkyl,2-acylglycerophosphocholines; LPCP: 1Z-alkenylglycerophosphocholines; PCP: 1-(1Z-alkenyl),2-acylglycerophosphocholines; oxPC: oxidized phosphatidylcholines; LPE: lyso-phosphatidylethanolamines.

**Fig. S5.**
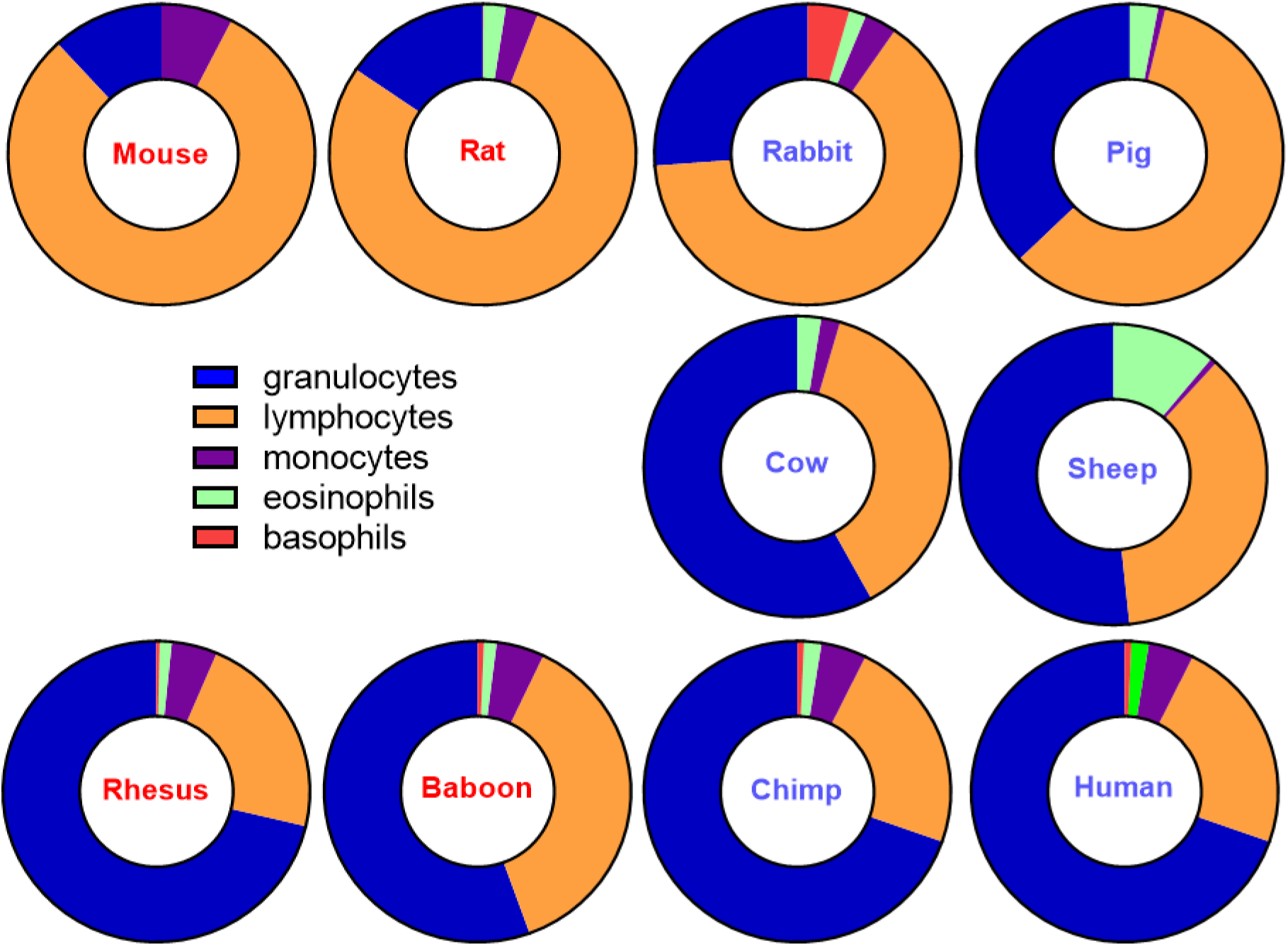
Blood differential counts obtained from the blood used in this study. Due to safety limitations, data was not obtained from the human subjects; established clinical reference values are given instead.

**Fig. S6.**
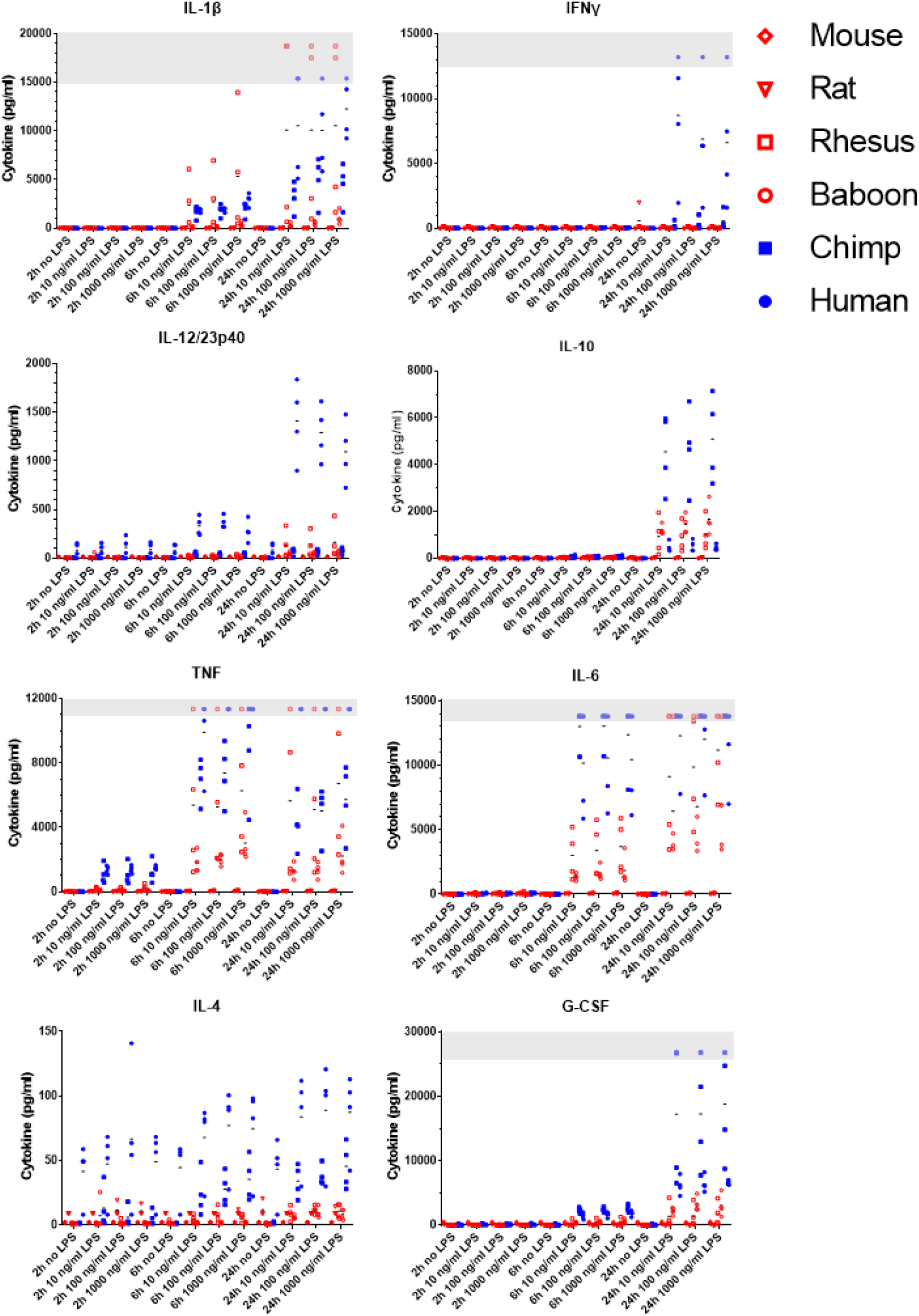
Cytokine release following LPS stimulation in the subset of species where antibodies were available, measured by Luminex. Black bars indicate mean values for each species. Grey bars indicate readings above quantifiable range.

**Fig. S7.**
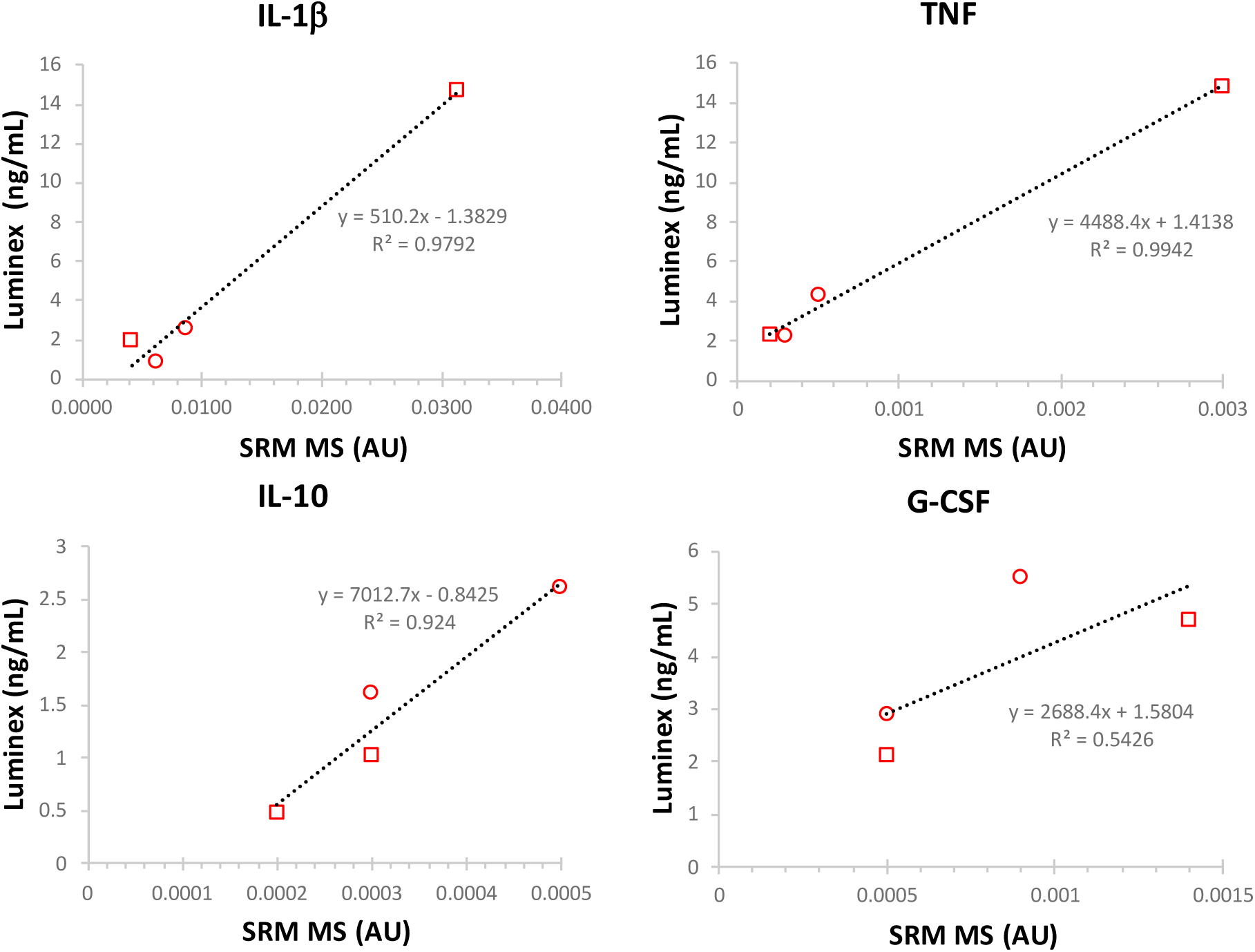
Validation of cytokine measurements. Concentrations of the indicated cytokines were established by sequence specific targeted SRM MS of unique peptides for two rhesus (squares) and two baboon (circles) plasma samples. Concentrations are expressed relative to peptide standard in arbitrary units (AU). Values are correlated with those obtained by Luminex for the same samples.

**Fig. S8.**
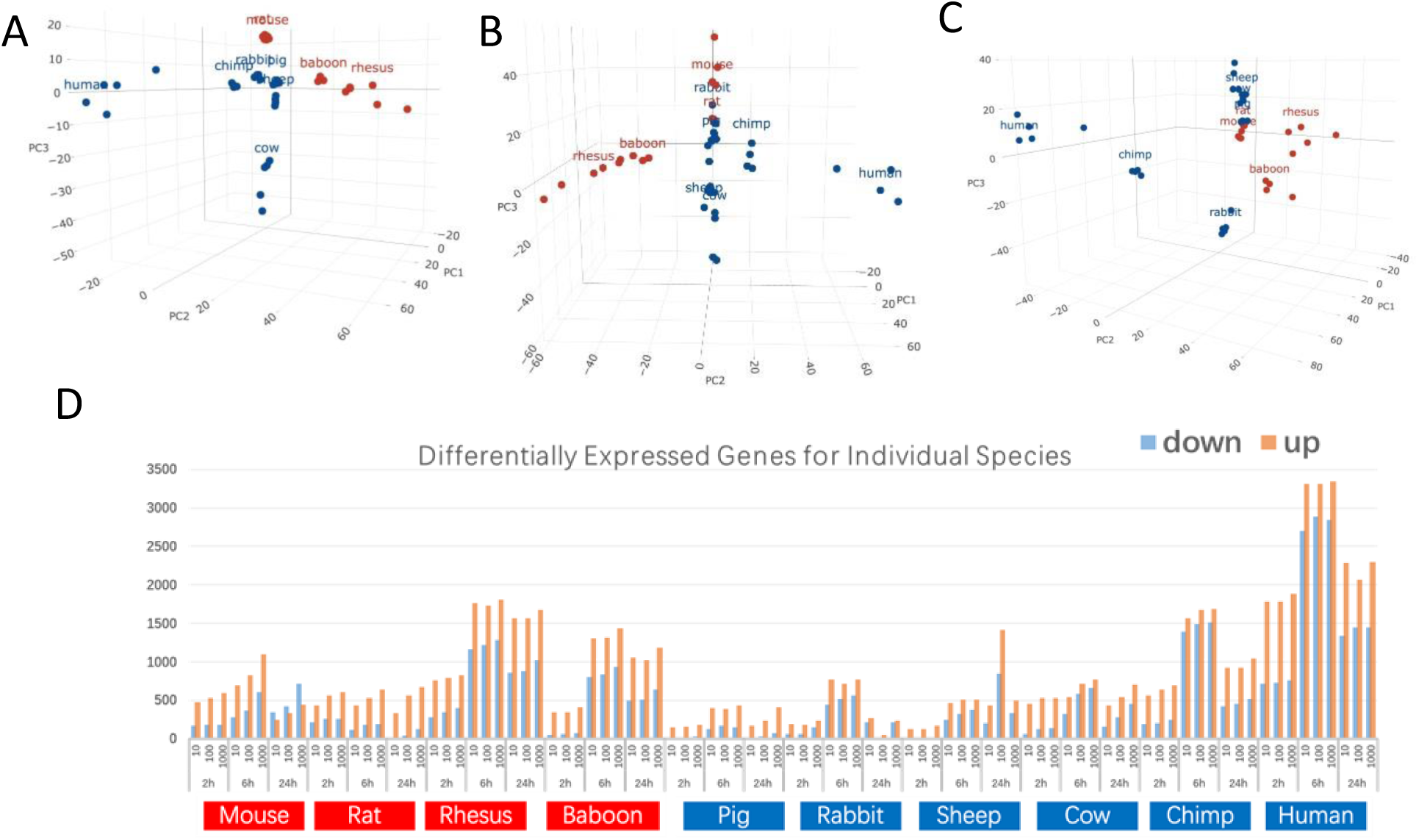
Gene expression responses of leukocytes to LPS stimulation. Principal component analysis showing overall distribution of fold change in individual mRNA abundance at A) 2h, B) 6h, or C) 24h following 10 ng/mL LPS stimulation. D) Number of genes significantly (2-fold change, FDR<0.05) changed relative to baseline at each time point after stimulation with 10, 100, or 1000 ng/mL LPS.

**Table S1.**
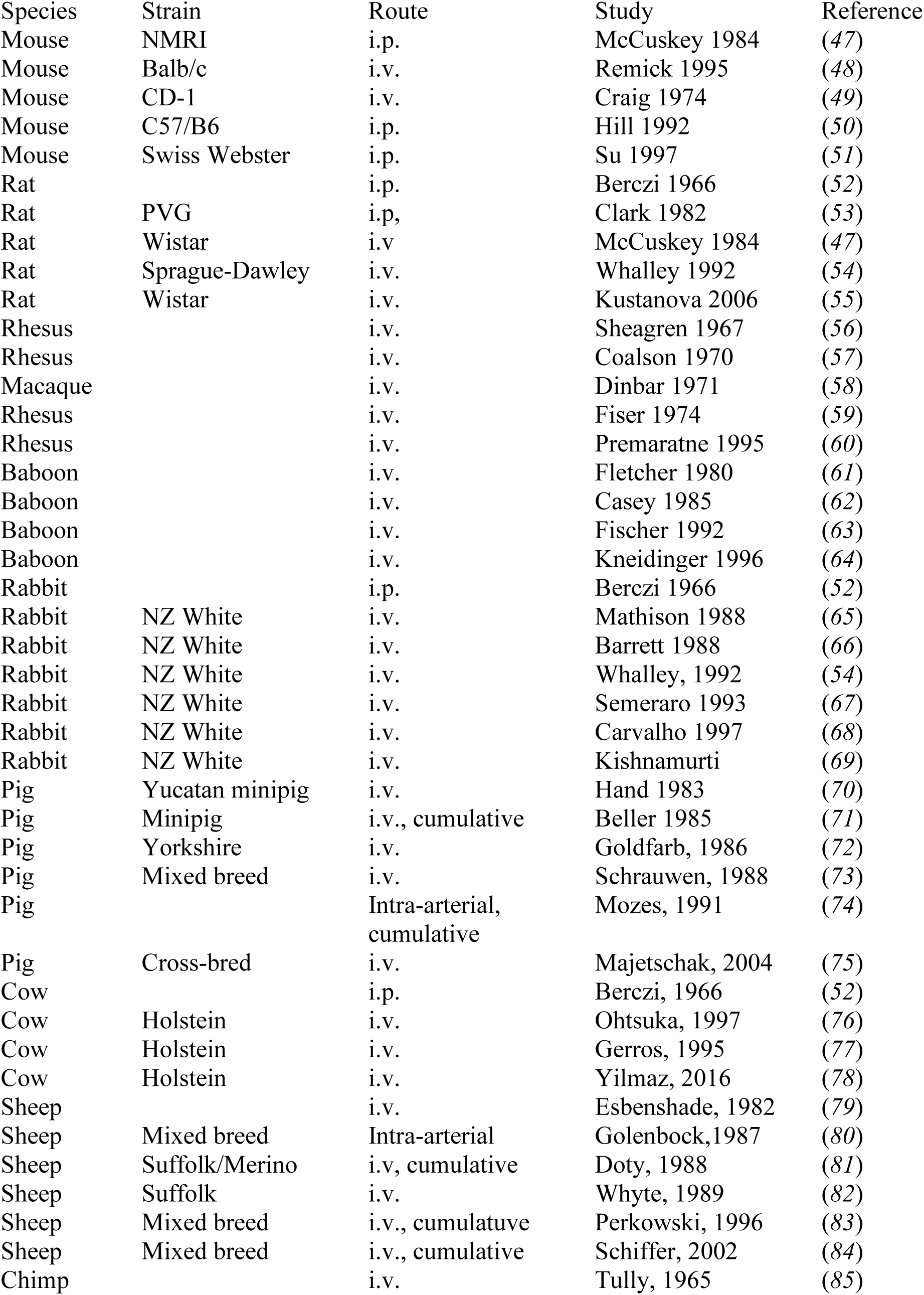
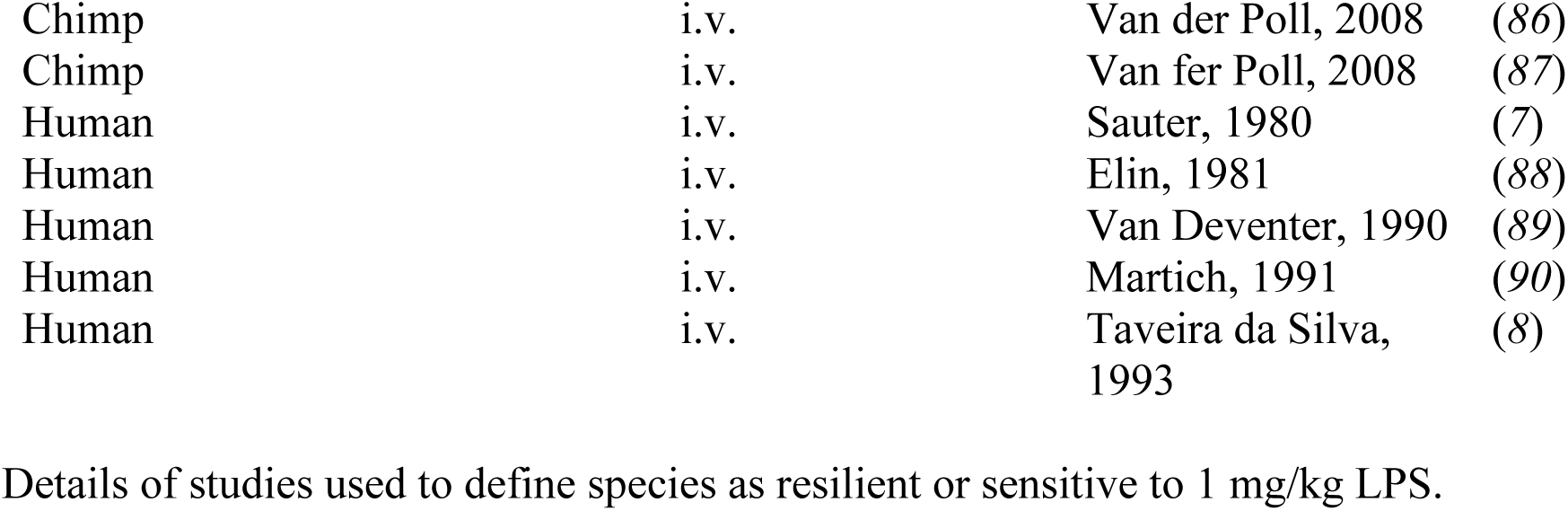
Details of studies used to define species as resilient or sensitive to 1 mg/kg LPS.

| Species | Strain | Route | Study | Reference |
| --- | --- | --- | --- | --- |
| Mouse | NMRI | i.p. | McCuskey 1984 | (47) |
| Mouse | Balb/c | i.v. | Remick 1995 | (48) |
| Mouse | CD-1 | i.v. | Craig 1974 | (49) |
| Mouse | C57/B6 | i.p. | Hill 1992 | (50) |
| Mouse | Swiss Webster | i.p. | Su 1997 | (51) |
| Rat |  | i.p. | Berczi 1966 | (52) |
| Rat | PVG | i.p. | Clark 1982 | (53) |
| Rat | Wistar | i.v. | McCuskey 1984 | (47) |
| Rat | Sprague-Dawley | i.v. | Whalley 1992 | (54) |
| Rat | Wistar | i.v. | Kustanova 2006 | (55) |
| Rhesus |  | i.v. | Sheagren 1967 | (56) |
| Rhesus |  | i.v. | Coalson 1970 | (57) |
| Macaque |  | i.v. | Dinbar 1971 | (58) |
| Rhesus |  | i.v. | Fiser 1974 | (59) |
| Rhesus |  | i.v. | Premaratne 1995 | (60) |
| Baboon |  | i.v. | Fletcher 1980 | (61) |
| Baboon |  | i.v. | Casey 1985 | (62) |
| Baboon |  | i.v. | Fischer 1992 | (63) |
| Baboon |  | i.v. | Kneidinger 1996 | (64) |
| Rabbit |  | i.p. | Berczi 1966 | (52) |
| Rabbit | NZ White | i.v. | Mathison 1988 | (65) |
| Rabbit | NZ White | i.v. | Barrett 1988 | (66) |
| Rabbit | NZ White | i.v. | Whalley, 1992 | (54) |
| Rabbit | NZ White | i.v. | Semeraro 1993 | (67) |
| Rabbit | NZ White | i.v. | Carvalho 1997 | (68) |
| Rabbit | NZ White | i.v. | Kishnamurti | (69) |
| Pig | Yucatan minipig | i.v. | Hand 1983 | (70) |
| Pig | Minipig | i.v., cumulative | Beller 1985 | (71) |
| Pig | Yorkshire | i.v. | Goldfarb, 1986 | (72) |
| Pig | Mixed breed | i.v. | Schrauwen, 1988 | (73) |
| Pig |  | Intra-arterial,<br>cumulative | Mozes, 1991 | (74) |
| Pig | Cross-bred | i.v. | Majetschak, 2004 | (75) |
| Cow |  | i.p. | Berczi, 1966 | (52) |
| Cow | Holstein | i.v. | Ohtsuka, 1997 | (76) |
| Cow | Holstein | i.v. | Gerros, 1995 | (77) |
| Cow | Holstein | i.v. | Yilmaz, 2016 | (78) |
| Sheep |  | i.v. | Esbenshade, 1982 | (79) |
| Sheep | Mixed breed | Intra-arterial | Golenbock, 1987 | (80) |
| Sheep | Suffolk/Merino | i.v, cumulative | Doty, 1988 | (81) |
| Sheep | Suffolk | i.v. | Whyte, 1989 | (82) |
| Sheep | Mixed breed | i.v., cumulative | Perkowski, 1996 | (83) |
| Sheep | Mixed breed | i.v., cumulative | Schiffer, 2002 | (84) |
| Chimp |  | i.v. | Tully, 1965 | (85) |

|  |  |  |  |
| --- | --- | --- | --- |
| Chimp | i.v. | Van der Poll, 2008 | (86) |
| Chimp | i.v. | Van fer Poll, 2008 | (87) |
| Human | i.v. | Sauter, 1980 | (7) |
| Human | i.v. | Elin, 1981 | (88) |
| Human | i.v. | Van Deventer, 1990 | (89) |
| Human | i.v. | Martich, 1991 | (90) |
| Human | i.v. | Taveira da Silva,<br>1993 | (8) |

## References and Notes

1. A. F. Read, A. L. Graham, L. Råberg, Animal Defenses against Infectious Agents: Is Damage Control More Important Than Pathogen Control. PLoS Biology 6, e1000004 (2008).

2. R. Medzhitov, D. S. Schneider, M. P. Soares, Disease Tolerance as a Defense Strategy. Science 335, 936–941 (2012).

3. L. Råberg, D. Sim, A. F. Read, Disentangling Genetic Variation for Resistance and Tolerance to Infectious Diseases in Animals. Science 318, 812–814 (2007).

4. J. S. Ayres, N. Freitag, D. S. Schneider, Identification of Drosophila Mutants Altering Defense of and Endurance to Listeria monocytogenes Infection. Genetics 178, 1807–1815 (2008).

5. A. M. Jamieson, L. Pasman, S. Yu, P. Gamradt, R. J. Homer, T. Decker, R. Medzhitov, Role of Tissue Protection in Lethal Respiratory Viral-Bacterial Coinfection. Science 340, 1230–1234 (2013).

6. S. F. Lowry, Human endotoxemia: a model for mechanistic insight and therapeutic targeting. *Shock (Augusta*, Ga.) 24 Suppl 1, 94–100 (2005).

7. C. Sauter, C. Wolfensberger, INTERFERON IN HUMAN SERUM AFTER INJECTION OF ENDOTOXIN. The Lancet 316, 852–853 (1980).

8. A. M. Taveira da Silva, H. C. Kaulbach, F. S. Chuidian, D. R. Lambert, A. F. Suffredini, R. L. Danner, Shock and Multiple-Organ Dysfunction after Self-Administration of Salmonella Endotoxin. New England Journal of Medicine 328, 1457–1460 (1993).

9. J.-M. Cavaillon, M. Adib-Conquy, Bench-to-bedside review: Endotoxin tolerance as a model of leukocyte reprogramming in sepsis. Critical Care 10, 233 (2006).

10. H. S. Warren, C. Fitting, E. Hoff, M. Adib-Conquy, L. Beasley-Topliffe, B. Tesini, X. Liang, C. Valentine, J. Hellman, D. Hayden, J. Cavaillon, Resilience to Bacterial Infection: Difference between Species Could Be Due to Proteins in Serum. The Journal of Infectious Diseases 201, 223–232 (2010).

11. S. Tanaka, D. Couret, A. Tran-Dinh, J. Duranteau, P. Montravers, A. Schwendeman, O. Meilhac, High-density lipoproteins during sepsis: from bench to bedside. Critical Care 24, 134 (2020).

12. V. Deretic, B. Levine, Autophagy balances inflammation in innate immunity. Autophagy 14, 243–251 (2018).

13. D. C. Bellissimo, C. Chen, Q. Zhu, S. Bagga, C.-T. Lee, B. He, G. B. Wertheim, M. Jordan, K. Tan, G. S. Worthen, D. G. Gilliland, N. A. Speck, Runx1 negatively regulates inflammatory cytokine production by neutrophils in response to Toll-like receptor signaling. Blood Advances 4, 1145–1158 (2020).

14. N. Suzuki, S. Suzuki, G. S. Duncan, D. G. Millar, T. Wada, C. Mirtsos, H. Takada, A. Wakeham, A. Itie, S. Li, J. M. Penninger, H. Wesche, P. S. Ohashi, T. W. Mak, W.-C. Yeh, Severe impairment of interleukin-1 and Toll-like receptor signalling in mice lacking IRAK-4. Nature 416, 750–754 (2002).

15. E. Carlsson, J. L. Ding, B. Byrne, SARM modulates MyD88-mediated TLR activation through BB-loop dependent TIR-TIR interactions. Biochimica et Biophysica Acta (BBA) - Molecular Cell Research 1863, 244–253 (2016).

16. I. Zanoni, F. Granucci, Role of CD14 in host protection against infections and in metabolism regulation. Frontiers in Cellular and Infection Microbiology 3 (2013).

17. R. S. Munford, J. P. Weiss, M. Lu, Biochemical transformation of bacterial lipopolysaccharides by acyloxyacyl hydrolase reduces host injury and promotes recovery. Journal of Biological Chemistry 295, 17842–17851 (2020).

18. W. H. Yang, D. M. Heithoff, P. V Aziz, B. Haslund-Gourley, J. S. Westman, S. Narisawa, A. B. Pinkerton, J. L. Millán, V. Nizet, M. J. Mahan, J. D. Marth, Accelerated Aging and Clearance of Host Anti-inflammatory Enzymes by Discrete Pathogens Fuels Sepsis. Cell Host & Microbe 24, 500–513.e5 (2018).

19. A. Liepelt, P. Hohlstein, H. Gussen, J. Xue, A. C. Aschenbrenner, T. Ulas, L. Buendgens, K. T. Warzecha, M. Bartneck, T. Luedde, C. Trautwein, J. L. Schultze, A. Koch, F. Tacke, Differential Gene Expression in Circulating CD14+ Monocytes Indicates the Prognosis of Critically Ill Patients with Sepsis. [Preprint] (2020). 10.3390/jcm9010127.

20. T. H. Flo, K. D. Smith, S. Sato, D. J. Rodriguez, M. A. Holmes, R. K. Strong, S. Akira, A. Aderem, Lipocalin 2 mediates an innate immune response to bacterial infection by sequestrating iron. Nature 432, 917–921 (2004).

21. F. Song, Y. Yi, C. Li, Y. Hu, J. Wang, D. E. Smith, H. Jiang, Regulation and biological role of the peptide/histidine transporter SLC15A3 in Toll-like receptor-mediated inflammatory responses in macrophage. Cell death & disease 9, 770 (2018).

22. Z. Zi, Z. Song, S. Zhang, Y. Ye, C. Li, M. Xu, Y. Zou, L. He, H. Zhu, Rubicon Deficiency Enhances Cardiac Autophagy and Protects Mice From Lipopolysaccharide-induced Lethality and Reduction in Stroke Volume. Journal of Cardiovascular Pharmacology 65 (2015).

23. P. Neubert, A. Homann, D. Wendelborn, A.-L. Bär, L. Krampert, M. Trum, A. Schröder, S. Ebner, A. Weichselbaum, V. Schatz, P. Linz, R. Veelken, J. Schulte-Schrepping, A. C. Aschenbrenner, T. Quast, C. Kurts, S. Geisberger, K. Kunzelmann, K. Hammer, K. J. Binger, J. Titze, D. N. Müller, W. Kolanus, J. L. Schultze, S. Wagner, J. Jantsch, NCX1 represents an ionic Na+ sensing mechanism in macrophages. PLOS Biology 18, e3000722 (2020).

24. C. Dostert, M. Grusdat, E. Letellier, D. Brenner, The TNF Family of Ligands and Receptors: Communication Modules in the Immune System and Beyond. Physiological Reviews 99, 115–160 (2019).

25. J. Seok, H. S. Warren, A. G. Cuenca, M. N. Mindrinos, H. V Baker, W. Xu, D. R. Richards, G. P. McDonald-Smith, H. Gao, L. Hennessy, C. C. Finnerty, C. M. López, S. Honari, E. E. Moore, J. P. Minei, J. Cuschieri, P. E. Bankey, J. L. Johnson, J. Sperry, A. B. Nathens, T. R. Billiar, M. A. West, M. G. Jeschke, M. B. Klein, R. L. Gamelli, N. S. Gibran, B. H. Brownstein, C. Miller-Graziano, S. E. Calvano, P. H. Mason, J. P. Cobb, L. G. Rahme, S. F. Lowry, R. V Maier, L. L. Moldawer, D. N. Herndon, R. W. Davis, W. Xiao, R. G. Tompkins, L. S. C. R. P. Inflammation and Host Response to Injury, Large Scale Collaborative Research Program, A. Abouhamze, U. G. J. Balis, D. G. Camp, A. K. De, B. G. Harbrecht, D. L. Hayden, A. Kaushal, G. E. O’Keefe, K. T. Kotz, W. Qian, D. A. Schoenfeld, M. B. Shapiro, G. M. Silver, R. D. Smith, J. D. Storey, R. Tibshirani, M. Toner, J. Wilhelmy, B. Wispelwey, W. H. Wong, Genomic responses in mouse models poorly mimic human inflammatory diseases. Proceedings of the National Academy of Sciences of the United States of America 110, 3507–12 (2013).

26. J. A. Rudbach, F. I. Akiya, R. J. Elin, H. D. Hochstein, M. K. Luoma, E. C. Milner, K. C. Milner, K. R. Thomas, Preparation and properties of a national reference endotoxin. Journal of clinical microbiology 3, 21–5 (1976).

27. R. Patro, S. M. Mount, C. Kingsford, Sailfish enables alignment-free isoform quantification from RNA-seq reads using lightweight algorithms. Nature biotechnology 32, 462–4 (2014).

28. C. Robert, M. Watson, Errors in RNA-Seq quantification affect genes of relevance to human disease. Genome Biology 16, 177 (2015).

29. M. E. Ritchie, B. Phipson, D. Wu, Y. Hu, C. W. Law, W. Shi, G. K. Smyth, limma powers differential expression analyses for RNA-sequencing and microarray studies. Nucleic Acids Research 43, e47–e47 (2015).

30. B. Chapman, R. Kirchner, L. Pantano, T. Khotiainsteva, M. De Smet, L. Beltrame, V. Saveliev, R. V. Guimera, S. Naumenko, J. Kern, C. Brueffer, G. Carrasco, M. Giovacchini, I. Sytchev, P. Tang, M. Ahdesmaki, S. Kanwal, J. J. Porter, S. Möller, V. Le, A. Coman, V. Svensson, bogdang989, M. Mistry, M. Edwards, J. Hammerbacher, B. Pedersen, P. Cock, apastore, S. Turner, bcbio/bcbio-nextgen: v1.1.9. doi: 10.5281/ZENODO.3564939 (2019).

31. S. E. Calvano, W. Xiao, D. R. Richards, R. M. Felciano, H. V. Baker, R. J. Cho, R. O. Chen, B. H. Brownstein, J. P. Cobb, S. K. Tschoeke, C. Miller-Graziano, L. L. Moldawer, M. N. Mindrinos, R. W. Davis, R. G. Tompkins, S. F. Lowry, Inflamm and Host Response to Injury Large Scale Collab. Res. Program, A network-based analysis of systemic inflammation in humans. Nature 437, 1032–1037 (2005).

32. A. Krämer, J. Green, J. Pollard, S. Tugendreich, Causal analysis approaches in Ingenuity Pathway Analysis. Bioinformatics 30, 523–530 (2014).

33. L. G. Jarsberg, K. Kedia, J. Wendler, A. T. Wright, P. D. Piehowski, M. A. Gritsenko, T. Shi, D. M. Lewinsohn, G. B. Sigal, M. H. Weiner, R. D. Smith, J. Keane, J. M. Jacobs, P. Nahid, Nutritional markers and proteome in patients undergoing treatment for pulmonary tuberculosis differ by geographic region. PLOS ONE 16, e0250586 (2021).

34. B. L. Deatherage Kaiser, J. M. Jacobs, A. A. Schepmoes, H. M. Brewer, B.-J. M. Webb-Robertson, S. Valtier, V. S. Bebarta, J. N. Adkins, Assessment of the Utility of the Oral Fluid and Plasma Proteomes for Hydrocodone Exposure. Journal of Medical Toxicology 16, 49–60 (2020).

35. O. Alcazar, L. F. Hernandez, E. S. Nakayasu, P. D. Piehowski, C. Ansong, M. H. Abdulreda, P. Buchwald, Longitudinal proteomics analysis in the immediate microenvironment of islet allografts during progression of rejection. Journal of Proteomics 223, 103826 (2020).

36. A. D. Polpitiya, W.-J. Qian, N. Jaitly, V. A. Petyuk, J. N. Adkins, D. G. Camp, G. A. Anderson, R. D. Smith, DAnTE: a statistical tool for quantitative analysis of -omics data. Bioinformatics 24, 1556–1558 (2008).

37. C. M. Henderson, T. Vaisar, A. N. Hoofnagle, “Isolating and Quantifying Plasma HDL Proteins by Sequential Density Gradient Ultracentrifugation and Targeted Proteomics” in Quantitative Proteomics by Mass Spectrometry (2016), pp. 105–120.

38. J. Folch, M. Lees, G. H. S. Stanley, A SIMPLE METHOD FOR THE ISOLATION AND PURIFICATION OF TOTAL LIPIDES FROM ANIMAL TISSUES. Journal of Biological Chemistry 226, 497–509 (1957).

39. E. S. Nakayasu, C. D. Nicora, A. C. Sims, K. E. Burnum-Johnson, Y.-M. Kim, J. E. Kyle, M. M. Matzke, A. K. Shukla, R. K. Chu, A. A. Schepmoes, J. M. Jacobs, R. S. Baric, B.-J. Webb-Robertson, R. D. Smith, T. O. Metz, MPLEx: a Robust and Universal Protocol for Single-Sample Integrative Proteomic, Metabolomic, and Lipidomic Analyses. mSystems 1 (2016).

40. J. E. Kyle, K. G. Stratton, E. M. Zink, Y.-M. Kim, K. J. Bloodsworth, M. E. Monroe, C. A. Bacino, N. A. Hanchard, R. A. Lewis, J. A. Rosenfeld, D. A. Scott, A. A. Tran, P. A. Ward, L. C. Burrage, G. D. Clark, M. E. Alejandro, J. E. Posey, M. F. Wangler, B. H. Lee, W. J. Craigen, H. J. Bellen, S. K. Nicholas, B. L. Bostwick, S. L. Samson, A. M. Goldman, P. M. Moretti, C. M. Eng, D. M. Muzny, J. P. Orengo, T. P. Vogel, S. R. Lalani, D. R. Murdock, M. S. Azamian, J. S. Orange, L. T. Emrick, S. U. Dhar, A. Balasubramanyam, L. Potocki, S. Yamamoto, Y. Yang, S. Chen, F. Jamal, L. Karaviti, R. Marom, S. A. Lincoln, C. A. Walsh, A. H. Beggs, L. H. Rodan, J. M. Stoler, G. T. Berry, L. A. Cobban, C. A. MacRae, J. B. Krier, E. K. Silverman, E. L. Fieg, R. L. Maas, J. Loscalzo, A. Aday, S. Korrick, D. B. Goldstein, N. Stong, J. A. Sullivan, R. C. Spillmann, L. D. M. Pena, Q. K.-G. Tan, N. M. Walley, Y. Jiang, A. McConkie-Rosell, K. Schoch, V. Shashi, H. Cope, I. A. Holm, I. S. Kohane, A. T. McCray, C. Esteves, K. LeBlanc, M. Might, E. Kelley, E. A. Worthey, D. C. Dorset, B. E. Boone, S. E. Levy, C. L. Birch, A. L. Jones, D. M. Brown, D. P. Bick, J. S. Newberry, J. Lazar, T. May, D. A. Sweetser, L. C. Briere, J. C. Pallais, C. M. Cooper, F. High, M. Walker, H. A. Colley, L. A. Mamounas, T. A. Manolio, E. A. Burke, R. A. Godfrey, C. A. Groden, W. A. Gahl, L. A. Wolfe, T. C. Markello, C. C. Lau, D. D. Draper, S. E. Gould, M. E. Nehrebecky, C. E. Wahl, G. F. Batzli, E. F. Macnamara, J. G. Dayal, D. J. Eckstein, J. J. Mulvihill, C. J. Tifft, T. K. Urv, A. L. Wise, J. L. Murphy, A. L. Gropman, E. M. Howerton, D. M. Krasnewich, J. M. Johnston, B. N. Pusey, D. R. Adams, V. V Maduro, M. C. V Malicdan, M. Davids, T. Estwick, D. Novacic, P. Sharma, C. Toro, G. Yu, B. Behnam, P. D’Souza, C. Ferreira, M. Morimoto, E. H. Baker, J. Yang, J.-P. F. Gourdine, M. Brush, M. Haendel, E. A. Ashley, J. A. Bernstein, J. B. Sampson, D. B. Zastrow, N. D. Friedman, J. D. Merker, C. E. McCormack, P. G. Fisher, J. M. Davidson, A. M. Dries, G. M. Enns, M. M. Majcherska, C. M. Reuter, D. M. Waggott, J. N. Kohler, T. R. Coakley, K. S. Smith, M. T. Wheeler, D. Bonner, L. Fernandez, J. Hom, Y. Huang, S. Marwaha, C. Zhao, J. A. Martínez-Agosto, E. C. Dell’Angelica, J. C. Papp, E. D. Douine, S. F. Nelson, M. G. Martin, C. G. S. Palmer, N. H. Parker, M. J. Butte, A. J. Yoon, S. K. Loo, B. L. Fogel, K. M. Dipple, J. S. Sinsheimer, P. Allard, H. Barseghyan, N. Dorrani, H. Lee, E. Vilain, A. Eskin, G. Renteria, R. Signer, J. Wan, A. Zheng, M. Westerfield, J. A. Phillips, J. D. Cogan, J. H. Newman, A. K. Robertson, R. Hamid, A. Bican, E. Brokamp, L. Duncan, M. Kozuira, L. Rives, L. Shakachite, K. M. Waters, B.-J. M. Webb-Robertson, D. M. Koeller, T. O. Metz, U. D. Network, A resource of lipidomics and metabolomics data from individuals with undiagnosed diseases. Scientific Data 8, 114 (2021).

41. J. E. Kyle, K. L. Crowell, C. P. Casey, G. M. Fujimoto, S. Kim, S. E. Dautel, R. D. Smith, S. H. Payne, T. O. Metz, LIQUID: an-open source software for identifying lipids in LC-MS/MS-based lipidomics data. Bioinformatics 33, 1744–1746 (2017).

42. T. Pluskal, S. Castillo, A. Villar-Briones, M. Orešič, MZmine 2: Modular framework for processing, visualizing, and analyzing mass spectrometry-based molecular profile data. BMC Bioinformatics 11, 395–395 (2010).

43. T. Shi, X. Sun, Y. Gao, T. L. Fillmore, A. A. Schepmoes, R. Zhao, J. He, R. J. Moore, J. Kagan, K. D. Rodland, T. Liu, A. Y. Liu, R. D. Smith, K. Tang, D. G. Camp, W.-J. Qian, Targeted Quantification of Low ng/mL Level Proteins in Human Serum without Immunoaffinity Depletion. Journal of Proteome Research 12, 3353–3361 (2013).

44. C. M. Nielson, K. S. Jones, R. F. Chun, J. M. Jacobs, Y. Wang, M. Hewison, J. S. Adams, C. M. Swanson, C. G. Lee, D. Vanderschueren, S. Pauwels, A. Prentice, R. D. Smith, T. Shi, Y. Gao, A. A. Schepmoes, J. M. Zmuda, J. Lapidus, J. A. Cauley, R. Bouillon, I. Schoenmakers, E. S. Orwoll, for the O. F. in M. (MrOS) R. Group, Free 25-Hydroxyvitamin D: Impact of Vitamin D Binding Protein Assays on Racial-Genotypic Associations. The Journal of Clinical Endocrinology & Metabolism 101, 2226–2234 (2016).

45. B. MacLean, D. M. Tomazela, N. Shulman, M. Chambers, G. L. Finney, B. Frewen, R. Kern, D. L. Tabb, D. C. Liebler, M. J. MacCoss, Skyline: an open source document editor for creating and analyzing targeted proteomics experiments. Bioinformatics 26, 966–968 (2010).

46. J. He, X. Sun, T. Shi, A. A. Schepmoes, T. L. Fillmore, V. A. Petyuk, F. Xie, R. Zhao, M. A. Gritsenko, F. Yang, N. Kitabayashi, S.-S. Chae, M. A. Rubin, J. Siddiqui, J. T. Wei, A. M. Chinnaiyan, W.-J. Qian, R. D. Smith, J. Kagan, S. Srivastava, K. D. Rodland, T. Liu, D. G. Camp II, Antibody-independent targeted quantification of TMPRSS2-ERG fusion protein products in prostate cancer. Molecular Oncology 8, 1169–1180 (2014).

47. R. S. McCuskey, P. A. McCuskey, R. Urbaschek, B. Urbaschek, Species differences in Kupffer cells and endotoxin sensitivity. Infection and immunity 45, 278–80 (1984).

48. D. Remick, P. Manohar, G. Bolgos, J. Rodriguez, L. Moldawer, G. Wollenberg, Blockade of tumor necrosis factor reduces lipopolysaccharide lethality, but not the lethality of cecal ligation and puncture. *Shock (Augusta*, Ga.) 4, 89–95 (1995).

49. W. A. Craig, J. H. Turner, C. M. Kunin, Prevention of the generalized Shwartzman reaction and endotoxin lethality by polymyxin B localized in tissues. Infection and immunity 10, 287–92 (1974).

50. M. R. Hill, R. E. McCallum, Identification of tumor necrosis factor as a transcriptional regulator of the phosphoenolpyruvate carboxykinase gene following endotoxin treatment of mice. Infection and immunity 60, 4040–50 (1992).

51. D. Su, R. I. Roth, M. Yoshida, J. Levin, Hemoglobin increases mortality from bacterial endotoxin. Infection and immunity 65, 1258–66 (1997).

52. I. Berczi, L. Bertók, T. Bereznai, COMPARATIVE STUDIES ON THE TOXICITY OF ESCHERICHIA COLI LIPOPOLYSACCHARIDE ENDOTOXIN IN VARIOUS ANIMAL SPECIES. Canadian Journal of Microbiology 12, 1070–1071 (1966).

53. I. A. Clark, Correlation between susceptibility to malaria and babesia parasites and to endotoxicity. Transactions of the Royal Society of Tropical Medicine and Hygiene 76, 4–7 (1982).

54. E. T. Whalley, J. A. Solomon, D. M. Modafferi, K. A. Bonham, J. C. Cheronis, CP-0127, a novel potent bradykinin antagonist, increases survival in rat and rabbit models of endotoxin shock. Agents and actions. Supplements 38 ( **Pt 3**), 413–20 (1992).

55. G. A. Kustanova, A. N. Murashev, V. L. Karpov, B. A. Margulis, I. V Guzhova, I. R. Prokhorenko, S. V Grachev, M. B. Evgen’ev, Exogenous heat shock protein 70 mediates sepsis manifestations and decreases the mortality rate in rats. Cell stress & chaperones 11, 276–86 (2006).

56. J. N. Sheagren, S. M. Wolff, N. R. Shulman, Febrile and hematologic responses of rhesus monkeys to bacterial endotoxin. The American journal of physiology 212, 884–90 (1967).

57. J. J. Coalson, L. B. Hinshaw, C. A. Guenter, The pulmonary ultrastructure in septic shock. Experimental and Molecular Pathology 12, 84–103 (1970).

58. A. Dinbar, S. I. Rapaport, M. J. Patch, W. Grant, E. W. Fonkalsrud, Hematologic effects of endotoxin on the macaque monkey. Surgery 70, 596–603 (1971).

59. R. H. Fiser, J. G. Denniston, W. R. Beisel, Endotoxemia in the Rhesus Monkey: Alterations in Host Lipid and Carbohydrate Metabolism. Pediatric Research 8, 13–17 (1974).

60. S. Premaratne, M. L. May, C. K. Nakasone, J. J. McNamara, Pharmacokinetics of endotoxin in a rhesus macaque septic shock model. The Journal of surgical research 59, 428–32 (1995).

61. J. R. Fletcher, P. W. Ramwell, Indomethacin treatment following baboon endotoxin shock improves survival. Advances in shock research 4, 103–11 (1980).

62. L. C. Casey, J. R. Fletcher, M. I. Zmudka, P. W. Ramwell, The role of thromboxane in primate endotoxin shock. Journal of Surgical Research 39, 140–149 (1985).

63. E. Fischer, M. A. Marano, K. J. Van Zee, C. S. Rock, A. S. Hawes, W. A. Thompson, L. DeForge, J. S. Kenney, D. G. Remick, D. C. Bloedow, Interleukin-1 receptor blockade improves survival and hemodynamic performance in Escherichia coli septic shock, but fails to alter host responses to sublethal endotoxemia. The Journal of Clinical Investigation 89, 1551–1557 (1992).

64. R. Kneidinger, S. Bahrami, H. Redl, G. Schlag, M. Robinson, Comparison of endothelial activation during endotoxic and posttraumatic conditions by serum analysis of soluble e-selectin in nonhuman primates. Journal of Laboratory and Clinical Medicine 128, 515–519 (1996).

65. J. C. Mathison, E. Wolfson, R. J. Ulevitch, Participation of tumor necrosis factor in the mediation of gram negative bacterial lipopolysaccharide-induced injury in rabbits. The Journal of clinical investigation 81, 1925–37 (1988).

66. T. J. Barrett, M. E. Potter, I. K. Wachsmuth, Bacterial endotoxin both enhances and inhibits the toxicity of Shiga-like toxin II in rabbits and mice. Infection and immunity 57, 3434–7 (1989).

67. N. Semeraro, R. Triggiani, P. Montemurro, L. G. Cavallo, M. Colucci, Enhanced endothelial tissue factor but normal thrombomodulin in endotoxin-treated rabbits. Thrombosis Research 71, 479–486 (1993).

68. G. L. Carvalho, G. Wakabayashi, M. Shimazu, T. Karahashi, M. Yoshida, S. Yamamoto, K. Matsushima, N. Mukaida, B. D. Clark, T. Takabayashi, C. T. Brandt, M. Kitajima, Anti-interleukin-8 monoclonal antibody reduces free radical production and improves hemodynamics and survival rate in endotoxic shock in rabbits. Surgery 122, 60–68 (1997).

69. C. Krishnamurti, A. J. Carter, P. Maglasang, J. R. Hess, M. A. Cutting, B. M. Alving, Cardiovascular toxicity of human cross-linked hemoglobin in a rabbit endotoxemia model. Critical care medicine 25, 1874–80 (1997).

70. M. S. Hand, M. J. Fettman, L. G. Chandrasena, J. L. Cleek, R. W. Phillips, Endotoxin dose. I. Hemodynamic, metabolic, and lethal consequences in Yucatan minipigs. The American journal of physiology 244, E385–98 (1983).

71. F. K. Beller, E. H. Schmidt, W. Holzgreve, J. Hauss, Septicemia during pregnancy: A study in different species of experimental animals. American Journal of Obstetrics and Gynecology 151, 967–975 (1985).

72. R. D. Goldfarb, L. M. Nightingale, P. Kish, P. B. Weber, D. J. Loegering, Left ventricular function during lethal and sublethal endotoxemia in swine. The American journal of physiology 251, H364–73 (1986).

73. E. Schrauwen, E. Cox, A. Houvenaghel, Escherichia coli sepsis and endotoxemia in conscious young pigs. Veterinary research communications 12, 295–303 (1988).

74. T. Mózes, S. Ben-Efraim, CornéJ. A. M. Tak, J. P. C. Heiligers, P. R. Saxena, I. L. Bonta, Serum levels of tumor necrosis factor determine the fatal or non-fatal course of endotoxic shock. Immunology Letters 27, 157–162 (1991).

75. M. Majetschak, S. M. Cohn, J. A. Nelson, E. H. Burton, U. Obertacke, K. G. Proctor, Effects of exogenous ubiquitin in lethal endotoxemia. Surgery 135, 536–43 (2004).

76. H. Ohtsuka, T. Higuchi, H. Matsuzawa, H. Sato, K. Takahashi, J. Takahashi, T. Yoshino, Inhibitory Effect of LPS-Induced Tumor Necrosis Factor in Calves Treated with Chlorpromazine or Pentoxifylline. Journal of Veterinary Medical Science 59, 1075–1077 (1997).

77. T. C. Gerros, S. D. Semrad, R. A. Proctor, Alterations in clinical, hematological and metabolic variables in bovine neonatal endotoxemia. Canadian journal of veterinary research = Revue canadienne de recherche veterinaire 59, 34–9 (1995).

78. Z. Yilmaz, O. Eralp Inan, M. Kocaturk, A. T. Baykal, O. Hacariz, I. Hatipoglu, A. Tvarijonaviciute, M. Cansev, J. Ceron, I. H. Ulus, Changes in serum proteins after endotoxin administration in healthy and choline-treated calves. BMC Veterinary Research 12, 210 (2016).

79. A. M. Esbenshade, J. H. Newman, P. M. Lams, H. Jolles, K. L. Brigham, Respiratory failure after endotoxin infusion in sheep: lung mechanics and lung fluid balance. Journal of Applied Physiology 53 (1982).

80. D. T. Golenbock, J. A. Will, C. R. Raetz, R. A. Proctor, Lipid X ameliorates pulmonary hypertension and protects sheep from death due to endotoxin. Infection and immunity 55, 2471–6 (1987).

81. S. Doty, L. Traber, D. Herndon, R. Kimura, H. Lubbesmeyer, S. Davenport, D. Traber, Beta endorphin, a vasoconstrictor during septic shock. The Journal of trauma 28, 131–9 (1988).

82. R. I. Whyte, H. S. Warren, E. Greene, M. L. Glennon, D. R. Robinson, W. M. Zapol, Tolerance to low-dose endotoxin in awake sheep. Journal of Applied Physiology 66 (1989).

83. S. Z. Perkowski, P. J. Sloane, J. A. Spath, T. H. Elsasser, J. K. Fisher, M. H. Gee, TNF-alpha and the pathophysiology of endotoxin-induced acute respiratory failure in sheep. Journal of applied physiology (Bethesda, Md. : 1985) 80, 564–73 (1996).

84. E. R. C. Schiffer, G. Reber, P. De Moerloose, D. R. Morel, Evaluation of unfractionated heparin and recombinant hirudin on survival in a sustained ovine endotoxin shock model. Critical care medicine 30, 2689–99 (2002).

85. J. G. Tully, S. Gaines, W. D. Tigertt, Studies on Infection and Immunity in Experimental Typhoid Fever VI. Response of Chimpanzees to Endotoxin and the Effect of Tolerance on Resistance to Oral Challenge. The Journal of Infectious Diseases 115, 445–455 (1965).

86. T. van der Poll, J. Jansen, D. van Leenen, M. von der Möhlen, M. Levi, H. ten Cate, H. Gallati, J. W. ten Cate, S. J. van Deventer, Release of soluble receptors for tumor necrosis factor in clinical sepsis and experimental endotoxemia. The Journal of infectious diseases 168, 955–60 (1993).

87. T. Van Der Poll, M. Levi, H. Ten Cate, J. Jansen, B. J. Biemond, B. L. Haagmans, A. Eerenberg, S. J. H. Van Deventer, C. E. Hack, J. W. Ten Cate, Effect of postponed treatment with an anti-tumour necrosis factor (TNF) F(ab’)2 fragment on endotoxin-induced cytokine and neutrophil responses in chimpanzees. Clinical and Experimental Immunology 100, 21–25 (2008).

88. R. J. Elin, S. M. Wolff, K. P. W. J. McAdam, L. Chedid, F. Audibert, C. Bernard, F. Oberling, Properties of Reference Escherichia coli Endotoxin and Its Phthalylated Derivative in Humans. Journal of Infectious Diseases 144, 329–336 (1981).

89. S. J. van Deventer, H. R. Büller, J. W. ten Cate, L. A. Aarden, C. E. Hack, A. Sturk, Experimental endotoxemia in humans: analysis of cytokine release and coagulation, fibrinolytic, and complement pathways. Blood 76, 2520–6 (1990).

90. G. D. Martich, R. L. Danner, M. Ceska, A. F. Suffredini, Detection of interleukin 8 and tumor necrosis factor in normal humans after intravenous endotoxin: the effect of antiinflammatory agents. Journal of Experimental Medicine 173 (1991).

